# Physical exercise increases the NODDI-derived neurite density index across white matter tracts in healthy older adults: results from the FIT4BRAIN randomized controlled trial

**DOI:** 10.64898/2026.08.31.26361810

**Authors:** Adriana L. Ruiz-Rizzo, Simon J. Schrenk, Stefan Brodoehl, Christiane Frahm, Christian Gaser, Marco Herbsleb, Christian Puta, Otto W. Witte, Kathrin Finke

## Abstract

Cross-sectional studies suggest associations between physical exercise and white matter in older adults, but evidence from randomized controlled trials is scarce. Neurite orientation dispersion and density imaging indices are biophysically informed metrics of white matter microstructure. Here, we tested whether a remotely delivered, 8-week multicomponent physical exercise intervention impacts neurite density (NDI) and orientation dispersion (ODI) across major white matter tracts in older adults. This secondary analysis of a randomized controlled trial included participants with available diffusion MRI data (n = 66; age: 66.4 ± 3.6 y; 43 females). Participants were randomized to a multicomponent exercise (PAG, n = 34) or an active control (CON, n = 32) intervention. Intervention effects on NDI/ODI were tested using linear mixed-effects and Bayesian multilevel models adjusted for age and sex. A significant Timepoint × Group interaction was observed for NDI (p = 0.003) but not for ODI (p = 0.785), further confirmed in Bayesian analyses for 22 white matter tracts, indicating a greater increase in NDI in the PAG. The standardized composite VO_2_max score increased from pre- to post-intervention within the PAG, although the Timepoint × Group interaction was not significant (p = 0.079). Across all participants, pre-to-post changes in mean NDI were positively correlated with changes in VO_2_max, but this association did not differ between groups. Our results indicate that white matter microstructure remains responsive to short-term, multicomponent physical exercise in older adults.

**Significance statement:** White matter microstructure deteriorates with aging. Given the ample health benefits of physical exercise, we investigated whether it can also improve white matter microstructural indices. In this randomized controlled trial, an 8-week remotely delivered multicomponent exercise program increased an MRI-based index of white matter tissue density across major tracts in older adults. These changes were not observed in an active control group. An index of white matter tissue orientation did not show intervention-related changes. Our findings provide experimental evidence that white matter microstructure remains sensitive and responsive to short-term multicomponent physical exercise in later life.

## 1. Introduction

Aging reduces white matter volume and compromises the integrity of myelinated axons (Groh and Simons, 2025). Physical exercise is associated with extensive health benefits (Myers et al., 2017; Fosstveit et al., 2024) and reduced dementia risk (Iso-Markku et al., 2022). Cross-sectional and longitudinal studies have consistently reported positive associations between physical activity, cardiorespiratory fitness, and white matter volume and integrity (Maleki et al., 2022), with higher cardiorespiratory fitness linked to preserved white matter in brain regions vulnerable to aging (Sexton et al., 2016; Faulkner et al., 2024). These findings suggest that physical exercise may help maintain white matter in older adults. However, randomized controlled trials (RCTs) are required to determine whether physical exercise directly modifies white matter microstructure in older adults.

To date, relatively few RCTs have examined *white* matter compared with *gray* matter outcomes following exercise (Silva et al., 2024). Given that white matter comprises about 50% of the total brain volume (Bullock et al., 2022) and is vulnerable to age-related metabolic, inflammatory, and vascular processes (Levit et al., 2020; Mendez Colmenares et al., 2023), understanding how physical exercise modifies white matter is critical to appraise the impact of physical exercise on brain health. Findings from the few existing RCTs are inconsistent. Some studies have not observed changes in fractional anisotropy (FA), a measure of diffusion directionality in white matter (Wassenaar et al., 2019), following aerobic exercise interventions (e.g., Voss et al., 2013; Fissler et al., 2017; Clark et al., 2019; Mendez Colmenares et al., 2021; Polk et al., 2023), while others report regionally restricted effects (e.g., Burzynska et al., 2017). One possible explanation for these inconsistencies lies in the use of FA, as FA may lack specificity in the presence of fiber dispersion or crossing (Kamiya et al., 2020). Consequently, the detection and interpretation of intervention-related changes in white matter microstructure have been limited.

More specific diffusion models may therefore be required. Neurite orientation dispersion and density imaging (NODDI) models water diffusion considering intracellular, extracellular, and cerebrospinal fluid compartments (Zhang et al., 2012). The intracellular compartment, the tissue of interest, captures the highly restricted diffusion *across* axons and dendrites and the unhindered diffusion *along* them (Zhang et al., 2012). The neurite density (NDI) and orientation dispersion (ODI) indices respectively represent the neurite tissue fraction and its spatial organization, thereby providing a biophysical characterization of white matter microstructure (Zhang et al., 2012; Chang et al., 2015). This distinction further allows testing whether potential intervention effects are reflected in increased density, decreased dispersion, or both. Most of the brain’s long-range connectivity is supported by ensembles of myelin-wrapped axonal projections organized in tracts (Bullock et al., 2022). Hence, a whole-brain approach can help capture the distributed effects of physical exercise on white matter.

Recently, the ‘FIT4BRAIN’ RCT (Schrenk et al., 2026) demonstrated that an 8-week remote multicomponent physical exercise intervention reduced BrainAGE, a global marker of structural brain aging (Saraei et al., 2025). However, global structural measures do not directly characterize white-matter microstructure. To our knowledge, no previous RCT has investigated whole-brain NODDI-derived white matter changes following a remotely delivered multicomponent exercise intervention in healthy older adults. The present study therefore complements these analyses by investigating diffusion-MRI-derived indices of neurite density and orientation dispersion across anatomically defined pathways.

Here, we examined whether the 8-week remote multicomponent physical exercise intervention improves NODDI indices compared with an active control intervention in healthy older adults. Based on previous findings, we hypothesized that there would be (i) widespread effects of physical exercise on NODDI-derived white-matter indices (NDI and ODI) across major white matter tracts; (ii) an association between intervention-induced changes in these indices and those in cardiorespiratory fitness; and (iii) an association between intervention changes in these indices and those in cognitive functions previously shown to improve following the intervention, specifically inhibition ability and visual memory (Schrenk et al., 2026).

## 2. Methods

### 2.1. Participants

Ninety-two healthy older adults (66.35 ± 3.64 years; n = 64 females) participated in the FIT4BRAIN RCT and were allocated to a physical activity group (“PAG,” n = 46) or an active control group receiving a progressive muscle relaxation intervention (“CON,” n = 46). The results of the full sample are reported in Schrenk et al. (2026). For the present secondary analyses, we included those with diffusion-weighted imaging (DWI) data both at baseline and after the eight-week intervention (n = 67). The reduced MRI sample was mainly due to the scanner becoming unavailable during the later phase of the study, preventing post-intervention DWI acquisition in some participants. Furthermore, one PAG participant was excluded due to excessive motion during either DWI session (i.e., above 3.5 interquartile ranges of the group’s median total motion index, tmi = 5.77). The final sample thus comprised 66 participants (mean age at baseline = 66.36 ± 3.62 years, range = 60-75; 43 females, 65.15%). All participants provided written informed consent before participation. The original study was approved by the ethics committee of Jena University Hospital (No. 2021-2345-BO) and conducted in accordance with the Declaration of Helsinki.

### 2.2. Experimental Design

FIT4BRAIN was a randomized, controlled physical exercise intervention trial with 1:1 allocation. The trial was prospectively registered at the German Clinical Trials Register (DRKS00028022). A detailed description of the study design has been reported previously (Schrenk et al., 2023, 2026). Briefly, participants in the PAG (n = 34) completed an eight-week multicomponent physical exercise program comprising aerobic, balance, and coordination exercises. The aerobic component consisted of medium-to high-intensity walking sessions, while balance and coordination exercise training included yoga lessons and juggling units, respectively. Participants in the CON (n = 32) completed an active control intervention consisting of progressive muscle relaxation and podcast listening matched in duration and frequency to the balance and coordination exercises in the PAG intervention. Adherence was monitored using actigraphy watches, which recorded exercise completion and heart rate during the intervention.

### 2.3. Assessment of cardiorespiratory fitness

Cardiorespiratory fitness assessment is described in detail by Schrenk et al. (2026). Briefly, three approaches were combined to estimate cardiorespiratory fitness: (a) a submaximal bicycle ergometer test; (b) the 6-minute walk test; and (c) a non-exercise-based prediction model. Each approach yielded an estimate of relative maximal oxygen uptake in *mL* · *kg^-1^* · *min^-1^*. The three estimated VO_2_max values were averaged at the participant level, and the resulting mean estimated VO_2_max was subsequently standardized using age-and sex-adjusted norms (Myers et al., 2017). We used the resulting standardized composite VO_2_max score in all analyses involving cardiorespiratory fitness. Consistent with the terminology used in the parent RCT (Schrenk et al., 2026), this standardized composite score is referred to as VO_2_max throughout the present manuscript.

### 2.4. MRI Data Acquisition

Magnetic Resonance Imaging (MRI) data were acquired at a Siemens MAGNETOM Trio 3T scanner (Siemens Healthcare, Erlangen, Germany) before the first session and after the last session of the respective intervention. All MRI data were acquired on the same scanner using the same head coil, acquisition protocol, and sequence parameters at both time points. Diffusion-weighted MRI data were acquired with a 2D spin-echo echo-planar imaging sequence. A total of 104 volumes were acquired with anterior-to-posterior phase-encoding direction, simultaneous multislice (96 interleaved slices), factor 4, and the following acquisition parameters: repetition time, TR = 3313 ms; time-to-echo, TE = 87.20 ms; flip angle, FA = 78°; voxel size = 1.5 mm^3^ isotropic; field-of-view, FOV = 210 mm. This primary dataset consisted of 16 volumes with b = 0 s/mm^2^ and 88 diffusion-weighted volumes acquired across 30 unique, non-collinear directions at three different b-shells (800, 1600, and 2500 s/mm^2^). To facilitate correction of susceptibility-induced geometric distortions, a reverse, posterior-to-anterior phase-encoding direction was also collected (104 volumes using the same diffusion-weighted sequence parameters). One high-resolution, T1-weighted anatomical volume was acquired with the following parameters: 3D MPRAGE sequence, GRAPPA acceleration mode with factor 2; TR = 2400 ms; TE = 2.22 ms; FA = 8°; voxel size = 0.8 mm^3^ isotropic; FOV = 256 mm; inversion time = 1000 ms; and 208 sagittal slices acquired with anterior-to-posterior phase-encoding direction.

### 2.5. MRI Data Preprocessing and Analysis

After DICOM-to-NIfTI conversion, diffusion MRI data were preprocessed following the TRACULA (TRActs Constrained by UnderLying Anatomy) pipeline (https://surfer.nmr.mgh.harvard.edu/fswiki/Tracula) (Yendiki et al., 2011) in FreeSurfer 7.4.1 (https://surfer.nmr.mgh.harvard.edu/fswiki/). The TRACULA pipeline required the preprocessing of the anatomical volumes first. Accordingly, the two high-resolution anatomical volumes (baseline and post-intervention) were preprocessed with FreeSurfer’s recon-all command for longitudinal data (https://surfer.nmr.mgh.harvard.edu/fswiki/LongitudinalProcessing). This preprocessing pipeline included the regular processing stream with recon-all for each image separately, followed by the creation of a within-subject template (‘base’) and the longitudinal processing scheme. FreeSurfer’s thalamic segmentation (Iglesias et al., 2018) was also conducted to improve the tract reconstruction around the thalamus. The TRACULA pipeline was conducted for all (anterior-to-posterior and posterior-to-anterior) images and both timepoints following the longitudinal/multiscan design (Yendiki et al., 2016) for the configuration file (https://surfer.nmr.mgh.harvard.edu/fswiki/dmrirc#Example4.3ALongitudinalstudy.2Cmultiplediffusionscanspersession). The preprocessing (‘trac-all -prep’ command) aimed to reduce B0 inhomogeneity and eddy-current distortions, compute total head motion during diffusion MRI (Yendiki et al., 2014), and perform intra-subject and inter-subject registration. Tensor fitting and anatomical prior computation for white-matter pathways (from a manually annotated set of training subjects) were also done during the preprocessing step. FSL’s bedpostX (https://fsl.fmrib.ox.ac.uk/fsl/docs/diffusion/bedpostx.html) was then run to fit the ball-and-stick model to the preprocessed diffusion MRI data (Behrens et al., 2007) using its GPU implementation ‘*bedpostx_gpu*’ (Hernández et al., 2013) on the high-performance computing cluster of Friedrich-Schiller-Universität Jena. Finally, the probability distributions of 42 major white matter tracts were estimated using the command ‘trac-all -path’ by simultaneously fitting each pathway’s shape to the ball-and-stick model results and the prior knowledge of pathway anatomy from the manually annotated set of training subjects in the TRACULA atlas. We then fitted the NODDI model to the diffusion MRI data using the AMICO (accelerated microstructure imaging via convex optimization) framework (Daducci et al., 2015) (https://github.com/daducci/AMICO/wiki) to obtain whole-brain, voxel-wise NDI and ODI values, both of which contribute to the directionality of water molecules within tissues (fractional anisotropy) (Zhang et al., 2012). NODDI distinguishes three types of microstructural tissue compartments: intracellular (the space of interest, bounded by neurite membranes that restrict water diffusion), extracellular (space *around* neurites, e.g., glial cells), and cerebrospinal fluid (Zhang et al., 2012). Finally, for each dataset, the NODDI maps resulting from AMICO were combined with the tract probability maps derived from TRACULA. More specifically, probability maps were thresholded at 20% of the maximum and normalized to form non-binary, probability-weighted tract ROIs. For each tract, a probability-weighted mean ODI and NDI (respectively) were computed and exported to table format for subsequent analyses.

Before proceeding with the statistical analyses, to optimize the number of comparisons used to test our hypotheses, we averaged NDI and ODI of the 32 left- and right-lateralized white-matter tracts (i.e., acoustic radiation, anterior thalamic radiation, arcuate fasciculus, cingulum bundles dorsal and ventral, corticospinal tract, extreme capsule, fornix, frontal aslant tract, inferior longitudinal fasciculus, middle longitudinal fasciculus, optic radiation, superior longitudinal fasciculi I, II, and III, and uncinate fasciculus). This procedure identified 26 white-matter tracts for NDI and 26 for ODI, which were then used for the group statistical analyses.

Total intracranial and white matter volumes, bilateral hippocampal volume, and white matter hypointensity volume were additionally extracted from the longitudinal FreeSurfer segmentations as conventional structural MRI measures.

### 2.6. Cognitive assessment

All participants of the FIT4BRAIN RCT underwent assessment with diverse neuropsychological tasks to derive the primary and secondary cognitive outcomes (Schrenk et al., 2023, 2026). Exploratory analyses to examine the intervention effect on the secondary cognitive outcomes showed group differences in the Rey-Osterrieth complex figure memory (Osterrieth, 1944) as well as in the Stroop Color-Word-Interference-Test (Bäumler, 1985) (Schrenk et al., 2026). Therefore, here we used these two measures to examine potential relationships with intervention-induced changes in NODDI indices.

### 2.7. Statistical Analyses

Baseline between-group comparisons for the intervention groups, across relevant continuous and categorical demographic, clinical, and neuroimaging variables, were examined using Welch’s two-sample t-test and Fisher’s exact test, respectively. Associations between relevant variables at baseline and between relevant variables’ pre-to-post intervention change (i.e., *Post value* − *Pre value*) were tested using Pearson correlations. A linear mixed-effects model assessed the effect of intervention on cardiorespiratory fitness (i.e., mean VO_2_max z-scores).

To test the effect of the intervention on NDI and ODI values, we implemented linear mixed-effects models using the *lme4* R package (Bates et al., 2015). The models for each outcome, *Y* ∈ {*NDI*, *ODI*}, included Timepoint (pre- and post-intervention), Group (PAG and CON), and the Timepoint × Group interaction as fixed factors, Age (continuous) and Sex (female and male) as covariates, and Subjects (n = 66) and Tracts (n = 26) as random factors. The model was specified as: *Y* ∼ *timepoint* * *group* + *sex* + *age* + (1 | *id*) + (1 | *tract*)). Note that the order in which the categorical fixed factors are mentioned reflects their coding, so the first mentioned level is the reference level. To aid interpretation, the relevant intervention effect is thus: (*CON_post_* − *CON_pre_*) − (*PAG_post_* − *PAG_pre_*). Both type III tests of all fixed effects and the corresponding model coefficients are reported.

To further examine tract-specific intervention effects, we additionally implemented Bayesian multilevel models. This approach allowed us to estimate tract-specific effects while accounting for the hierarchical structure of the data. More specifically, we estimated the models using Markov chain Monte Carlo sampling (four chains; 12,000 iterations each, including 2,000 warm-up iterations) with the *brms* R package (v. 2.22.0; Bürkner, 2017). For this purpose, we explicitly modeled tract-wise random intercepts and slopes for the interaction for each outcome, *Y* ∈ {*NDI*, *ODI*} using the model specification: *Y* ∼ *timepoint* * *group* + *sex* + *age* + (1 + *timepoint* * *group* | *tract*) + (1 | *id*). Within this framework, model estimates were deemed to support our hypotheses if the Bayes factor (BF) ≥ 10 or the posterior probability of direction (pd) ≥ 95%. To obtain tract-specific estimates of the interaction term, the random slopes were combined with the population-level (fixed) interaction term, and significance was assessed using 95% credible intervals.

A mixed-effects model assessed the effect of intervention on cardiorespiratory fitness (i.e., mean VO_2_max z-scores). NODDI indices were averaged across tracts to compute Pearson’s correlations between NODDI indices, VO_2_max, and cognitive outcomes. Group differences in the correlation coefficients were tested with Fisher’s r-to-z transformation.

Sensitivity analyses were conducted to assess the effect of potentially differing scanning conditions at the two time points (e.g., head motion). More specifically, first, we examined main and interaction effects in head motion (tmi). Second, tmi was added as a covariate in the linear mixed-effects models of NDI and ODI. Last, a random term representing a varying intercept for each unique participant-tract combination (i.e., (1 | *id*: *tract*)) was added to the models of NDI and ODI to additionally account for the nested structure of tracts *within* participants, thereby capturing pre-post similarities of the same tract within the same participant.

Finally, between-group differences in exercise intensity, the average and maximum heart rate (in beats per minute) throughout the intervention, as recorded by the activity tracker, were estimated using analysis of covariance (ANCOVA) with Group as a between-subjects factor and Age and Sex as covariates.

All results were considered statistically significant at α = 0.05. False discovery rate (FDR) correction according to Benjamini and Hochberg was applied when multiple comparisons were conducted. Statistical analyses and visualizations were performed using R (version 4.5.0; R Core Team, 2025) in RStudio (version 2025.5.0.496; Posit team, 2025), and Python 3.12 in Google Colab (heart rate analysis). Analysis scripts, including the packages used, are available at https://github.com/alruizzo/F4B-WM. The data for the main analyses can be accessed before manuscript publication at peer-review-link.

## 3. Results

### 3.1. Descriptive results and group differences at baseline

Descriptive statistics on the demographic, health-related, and neuroimaging variables at baseline are listed in Table 1. No baseline differences were observed between the PAG and the CON for any variable. Both groups adhered comparably well to their corresponding interventions (PAG: 26.8 ± 3.1 recorded units versus CON: 24.9 ± 4.9 units, maximum: 29 units; t = -1.78, p = 0.080). As expected, actigraphy-derived heart rate measures during the intervention differed between groups (see Table 1-1).

**Table 1.**
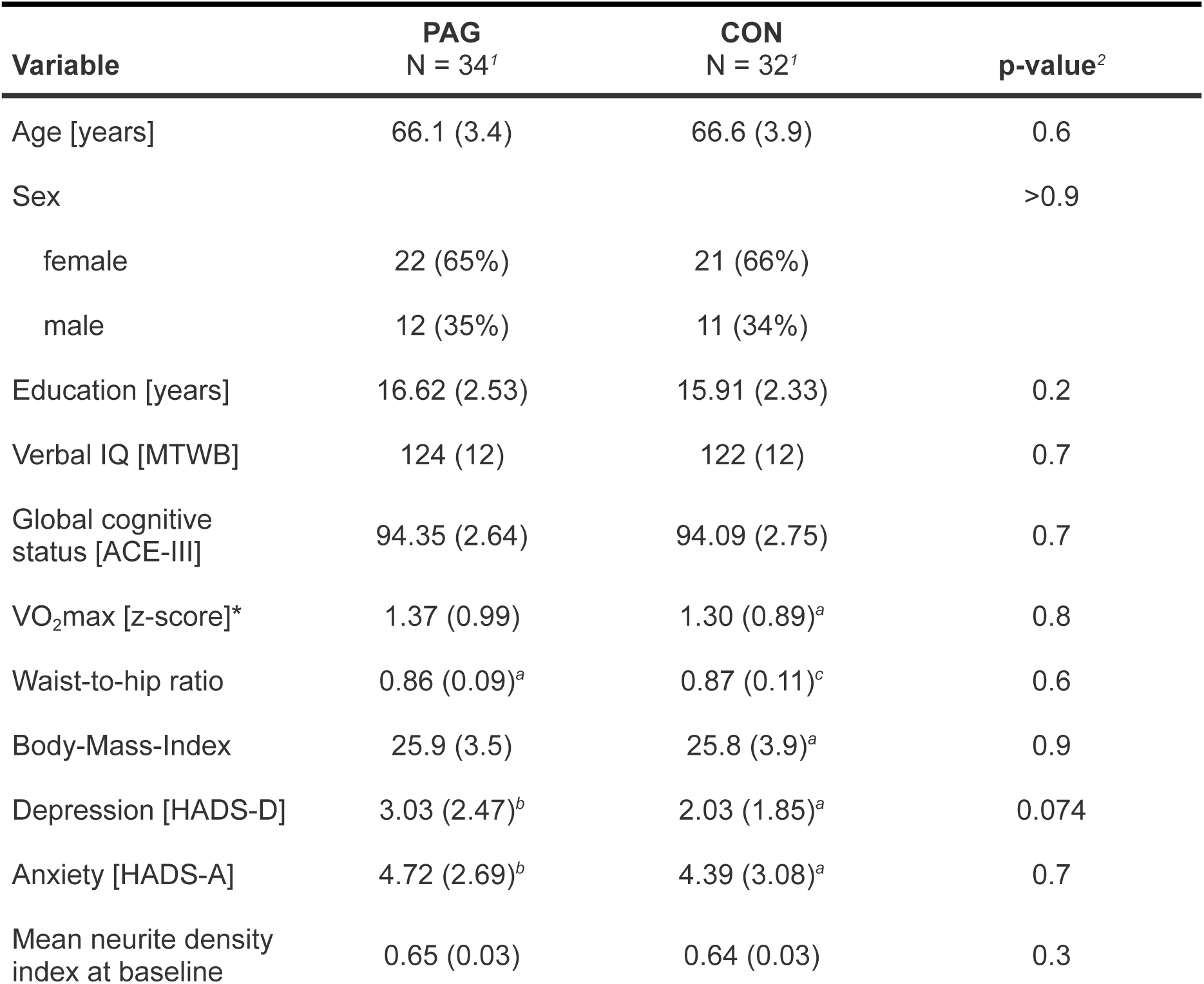
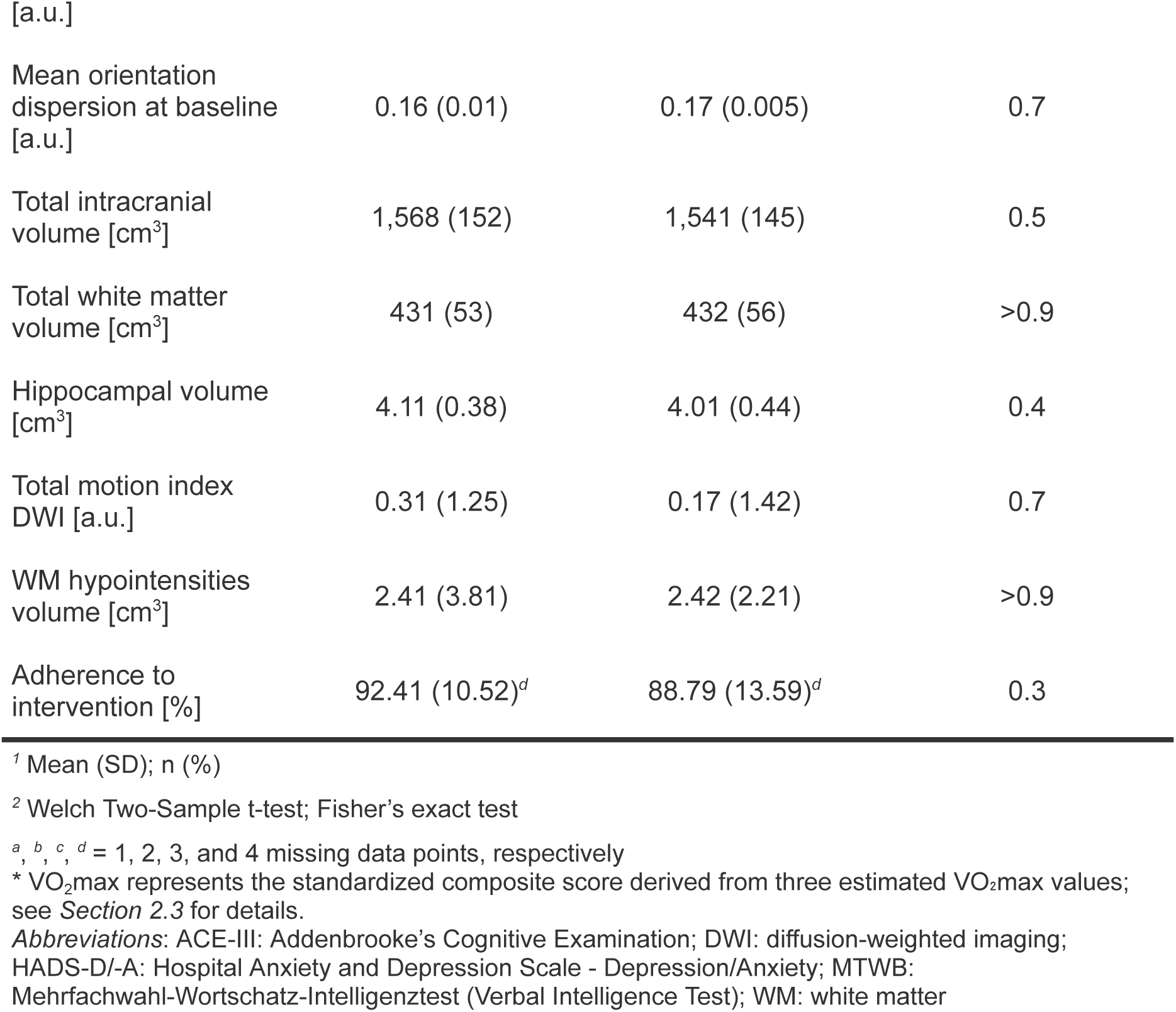
Baseline demographic, health-related, and neuroimaging variables for the active (‘PAG’) and control (‘CON’) intervention groups.

| Variable | PAG<br>N = 34<sup>1</sup> | CON<br>N = 32<sup>1</sup> | p-value<sup>2</sup> |
| --- | --- | --- | --- |
| Age [years] | 66.1 (3.4) | 66.6 (3.9) | 0.6 |
| Sex |  |  | >0.9 |
| female | 22 (65%) | 21 (66%) |  |
| male | 12 (35%) | 11 (34%) |  |
| Education [years] | 16.62 (2.53) | 15.91 (2.33) | 0.2 |
| Verbal IQ [MTWB] | 124 (12) | 122 (12) | 0.7 |
| Global cognitive status [ACE-III] | 94.35 (2.64) | 94.09 (2.75) | 0.7 |
| VO<sub>2</sub>max [z-score]* | 1.37 (0.99) | 1.30 (0.89)<sup>a</sup> | 0.8 |
| Waist-to-hip ratio | 0.86 (0.09)<sup>a</sup> | 0.87 (0.11)<sup>c</sup> | 0.6 |
| Body-Mass-Index | 25.9 (3.5) | 25.8 (3.9)<sup>a</sup> | 0.9 |
| Depression [HADS-D] | 3.03 (2.47)<sup>b</sup> | 2.03 (1.85)<sup>a</sup> | 0.074 |
| Anxiety [HADS-A] | 4.72 (2.69)<sup>b</sup> | 4.39 (3.08)<sup>a</sup> | 0.7 |
| Mean neurite density index at baseline | 0.65 (0.03) | 0.64 (0.03) | 0.3 |
| [a.u.] |  |  |  |
| Mean orientation dispersion at baseline [a.u.] | 0.16 (0.01) | 0.17 (0.005) | 0.7 |
| Total intracranial volume [cm<sup>3</sup>] | 1,568 (152) | 1,541 (145) | 0.5 |
| Total white matter volume [cm<sup>3</sup>] | 431 (53) | 432 (56) | >0.9 |
| Hippocampal volume [cm<sup>3</sup>] | 4.11 (0.38) | 4.01 (0.44) | 0.4 |
| Total motion index DWI [a.u.] | 0.31 (1.25) | 0.17 (1.42) | 0.7 |
| WM hypointensities volume [cm<sup>3</sup>] | 2.41 (3.81) | 2.42 (2.21) | >0.9 |
| Adherence to intervention [%] | 92.41 (10.52)<sup>d</sup> | 88.79 (13.59)<sup>d</sup> | 0.3 |

<sup>1</sup> Mean (SD); n (%)
<sup>2</sup> Welch Two-Sample t-test; Fisher's exact test
<sup>a</sup>, <sup>b</sup>, <sup>c</sup>, <sup>d</sup> = 1, 2, 3, and 4 missing data points, respectively
\* VO<sub>2</sub>max represents the standardized composite score derived from three estimated VO<sub>2</sub>max values; see Section 2.3 for details.
*Abbreviations:* ACE-III: Addenbrooke's Cognitive Examination; DWI: diffusion-weighted imaging; HADS-D/-A: Hospital Anxiety and Depression Scale - Depression/Anxiety; MTWB: Mehrfachwahl-Wortschatz-Intelligenztest (Verbal Intelligence Test); WM: white matter

The 26 major white matter tracts identified with TRACULA, from which NDI and ODI values were derived, are shown in Figure 1. NDI and ODI values did not differ between the intervention groups at baseline after FDR correction (Figures 1-1 and 1-2). Correlations between NDI and ODI values across tracts are provided in the Extended Data.

**Figure 1.**
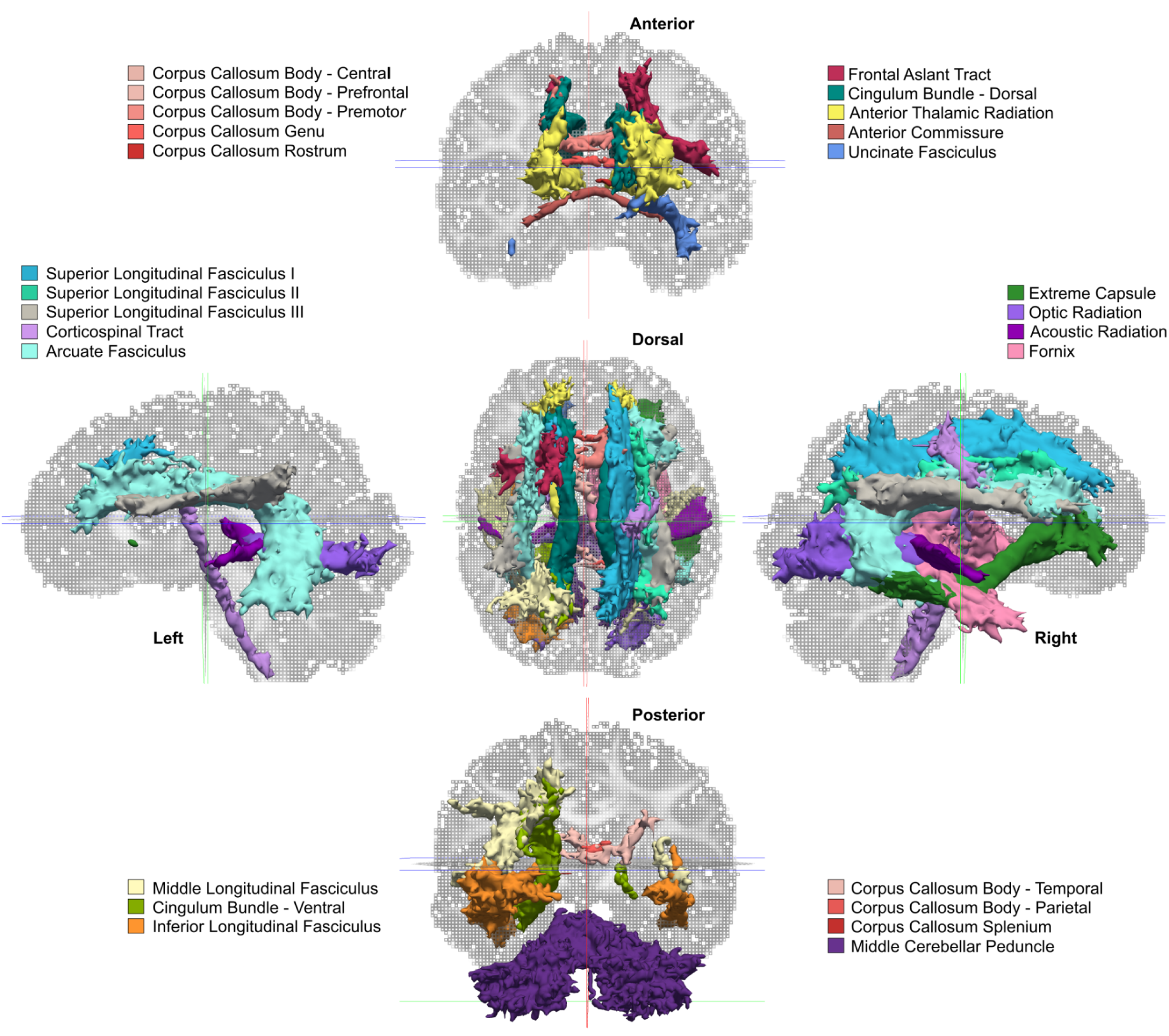
Major white-matter tracts reconstructed with TRACULA (TRActs Constrained by UnderLying Anatomy) in a representative study participant. Visualization of the probabilistic 42 white-matter pathways used to derive neurite density (NDI) and orientation dispersion (ODI) indices. Tracts are overlaid on the participant’s fractional anisotropy image. For clarity, bilateral tracts are mentioned only once.

### 3.2. Effect of intervention on cardiorespiratory fitness

Pre- and post-intervention VO_2_max values are plotted in Figure 2. The corresponding model estimates are presented in Table 2. Type III analyses revealed that the Timepoint × Group interaction (*F*_(1, 60.1)_ = 3.18, p = 0.079) was not significant. Follow-up contrasts of the estimated marginal means indicated an increase in VO_2_max in the PAG (estimate of the Post – Pre difference = 0.09, SE = 0.04, t_(60.1)_ = 2.50, p = 0.030), whereas no statistically significant pre-to-post change was detected in the CON (Post – Pre difference = -0.002, SE = 0.04, t_(60.1)_ = -0.06, p = 0.950). The between-group difference at post-intervention was also not significant (PAG – CON difference = 0.15, SE = 0.21, t_(63.2)_ = 0.72, p = 0.951). A main effect of sex was observed (see Table 2), with female participants showing higher VO_2_max values relative to their sex-specific reference values than male participants showed relative to theirs.

**Figure 2.**
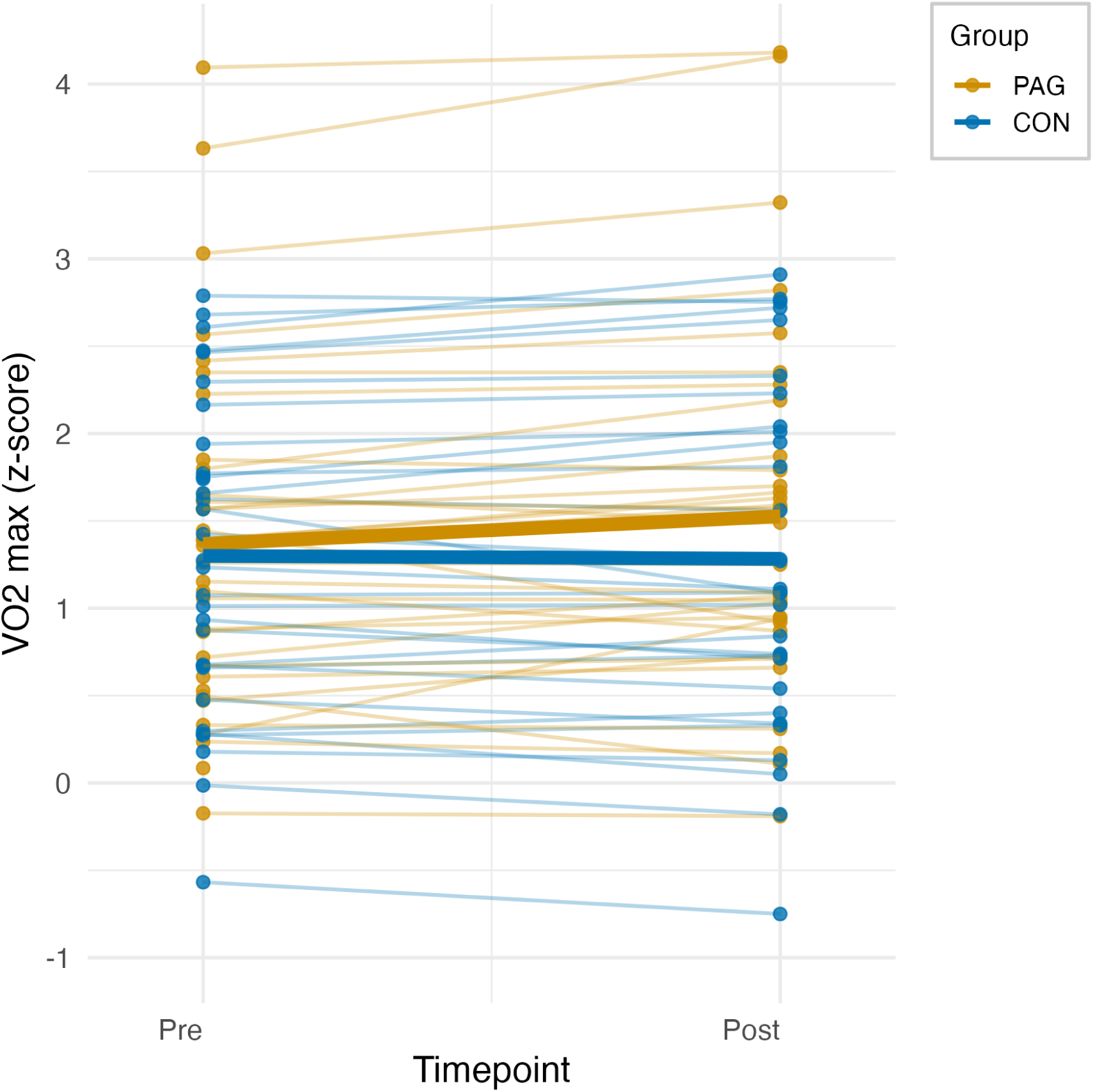
Spaghetti plot of cardiorespiratory fitness (mean VO_2_max z-score) pre-and post-intervention. Individual values are plotted as dots, color-coded by group: physical exercise (“PAG”) in dark yellow and control intervention (“CON”) in blue. The thicker lines represent the group means. The Timepoint × Group interaction was not statistically significant (p = 0.079, 95% CI [-0.20, 0.01]; see Table 2).

**Table 2.**
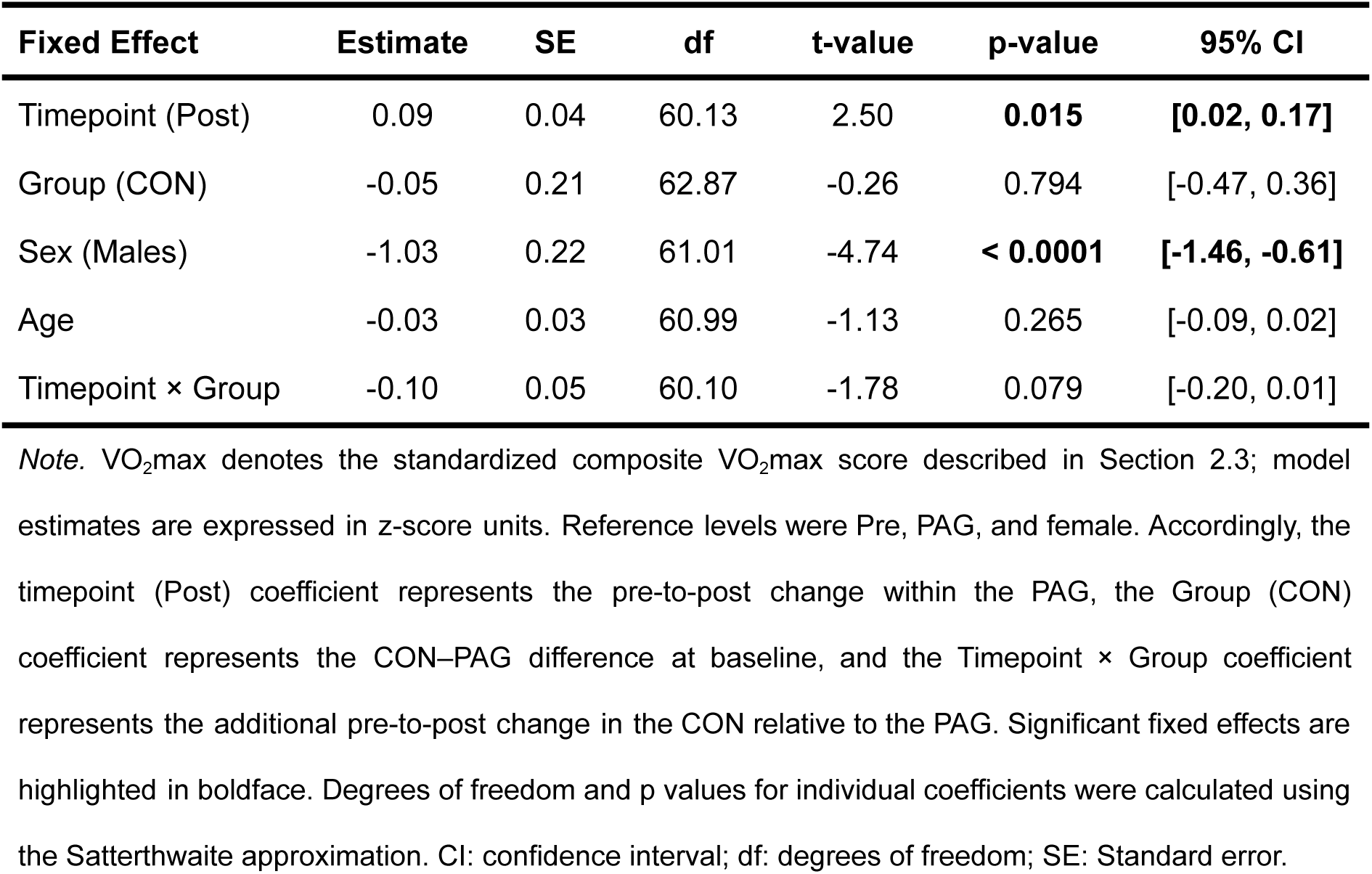
Results of the mixed-effects model for mean VO_2_max (standardized composite score)

### 3.3. Effect of intervention type on neurite density and orientation dispersion

The Type III analysis of a mixed-effects model revealed a significant Timepoint × Group interaction for NDI (*F*_(1, 3339)_ = 8.58, p = 0.003). No main effects of Group (*F*_(1, 62)_ = 2.26, p = 0.138) or Timepoint (*F*_(1, 3339)_ = 0.12, p = 0.728) were observed. In addition, significant effects of Sex (*F*_(1, 62)_ = 10.46,p = 0.002) and Age (*F*_(1, 62)_ = 8.32,p = 0.005) were found. The fixed-effect parameter estimates are provided in Table 3.

**Table 3.** Results of the mixed-effects model on NDI.

| Fixed Effect | Estimate | SE | df | t-value | p-value | 95% CI |
| --- | --- | --- | --- | --- | --- | --- |
| Timepoint (Post) | 0.003 | 0.001 | 3339 | 2.35 | **0.019** | **[0.0005, 0.005]** |
| Group (CON) | -0.006 | 0.006 | 64.5 | -1.08 | 0.285 | [-0.018, 0.005] |
| Sex (Males) | 0.02 | 0.01 | 62 | 3.23 | **0.002** | **[0.01, 0.03]** |
| Age | -0.002 | 0.001 | 62 | -2.88 | **0.005** | **[-0.004, -0.001]** |
| Timepoint × Group | -0.005 | 0.002 | 3339 | -2.93 | **0.003** | **[-0.01, -0.002]** |

*Note.* The reference levels for the first three variables are Pre, PAG, and females, from top to bottom. Model: $NDI \sim \text{timepoint} * \text{group} + \text{sex} + \text{age} + (1 | \text{id}) + (1 | \text{tract})$ . The timepoint estimate represents the pre-to-post change in the PAG group. Significant fixed effects are highlighted in boldface. Degrees of freedom and p-values for individual coefficients were calculated using the Satterthwaite approximation. CI: confidence interval; df: degrees of freedom; SE: Standard error.

Follow-up tests indicated a significant increase in NDI from pre- to post-intervention in the PAG (Δ = 0.003, SE = 0.001, t_(3339)_ = 2.35, p = 0.037, 95% CI [0.0001, 0.005]), but not in the CON (Δ = -0.002, SE = 0.001, t_(3339)_ = -1.80, p = 0.072, 95% CI [-0.005, 0.0005]). Post-intervention NDI tended to be higher in the PAG than CON, although the difference did not reach significance (Δ = 0.01, SE = 0.006, t_(64.5)_ = 1.90, p = 0.125, 95% CI [-0.002, 0.025]). For ODI, neither the Timepoint × Group interaction nor any of the main effects reached significance (all *p*-values > 0.388; Table 4).

**Table 4.**
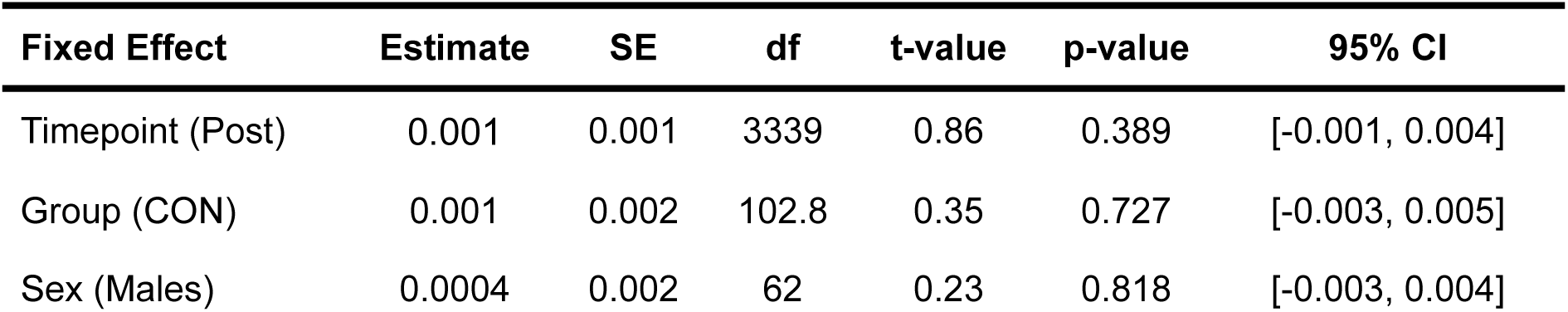
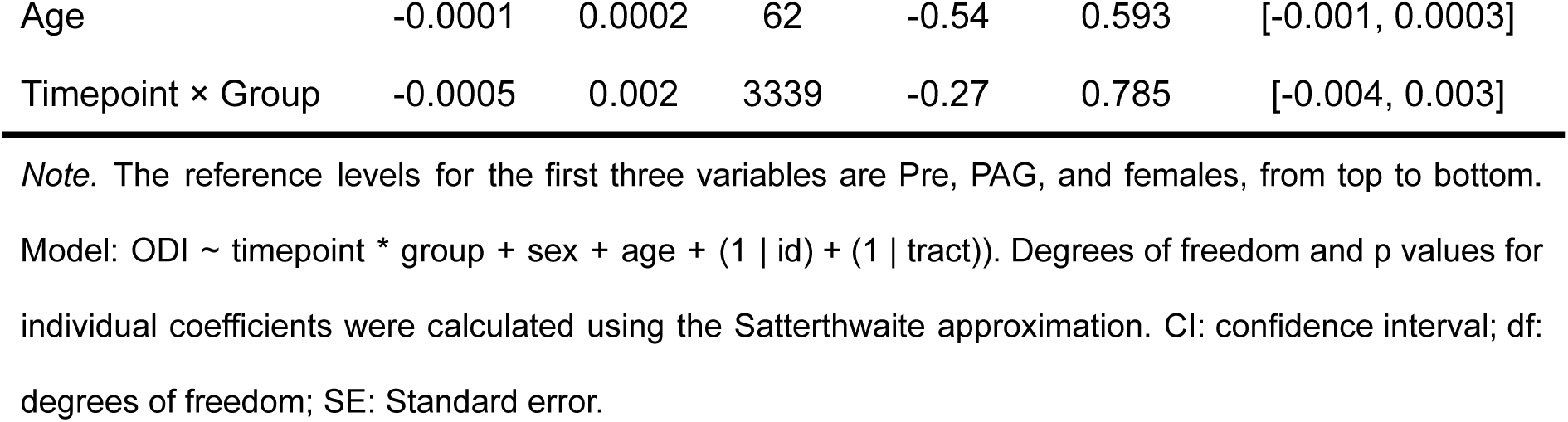
Results of the mixed-effects model on ODI.

| Fixed Effect | Estimate | SE | df | t-value | p-value | 95% CI |
| --- | --- | --- | --- | --- | --- | --- |
| Timepoint (Post) | 0.001 | 0.001 | 3339 | 0.86 | 0.389 | [-0.001, 0.004] |
| Group (CON) | 0.001 | 0.002 | 102.8 | 0.35 | 0.727 | [-0.003, 0.005] |
| Sex (Males) | 0.0004 | 0.002 | 62 | 0.23 | 0.818 | [-0.003, 0.004] |
| Age | -0.0001 | 0.0002 | 62 | -0.54 | 0.593 | [-0.001, 0.0003] |
| Timepoint × Group | -0.0005 | 0.002 | 3339 | -0.27 | 0.785 | [-0.004, 0.003] |

*Note.* The reference levels for the first three variables are Pre, PAG, and females, from top to bottom.
Model: $ODI \sim \text{timepoint} * \text{group} + \text{sex} + \text{age} + (1 | \text{id}) + (1 | \text{tract})$ . Degrees of freedom and p values for individual coefficients were calculated using the Satterthwaite approximation. CI: confidence interval; df: degrees of freedom; SE: Standard error.

### 3.4. Effect of intervention type on neurite density and orientation dispersion on individual major white-matter tracts

Consistent with the mixed-effects model, the Bayesian multilevel model on NDI values revealed strong evidence for a Timepoint × Group interaction (Bayes factor, BF = 21.11; estimate: -0.005, 95% credible interval [-0.01, -0.002], posterior probability of direction, pd = 99.9%; see Table 3-1). At the tract level, 22 of the 26 major white matter tracts showed interaction estimates whose 95% credible intervals did not include zero (Figure 3), indicating a widespread effect across white matter. Non-significant effects were observed for the middle cerebellar peduncle, the corticospinal tract, and the corpus callosum (body-central and splenium).

**Figure 3.**
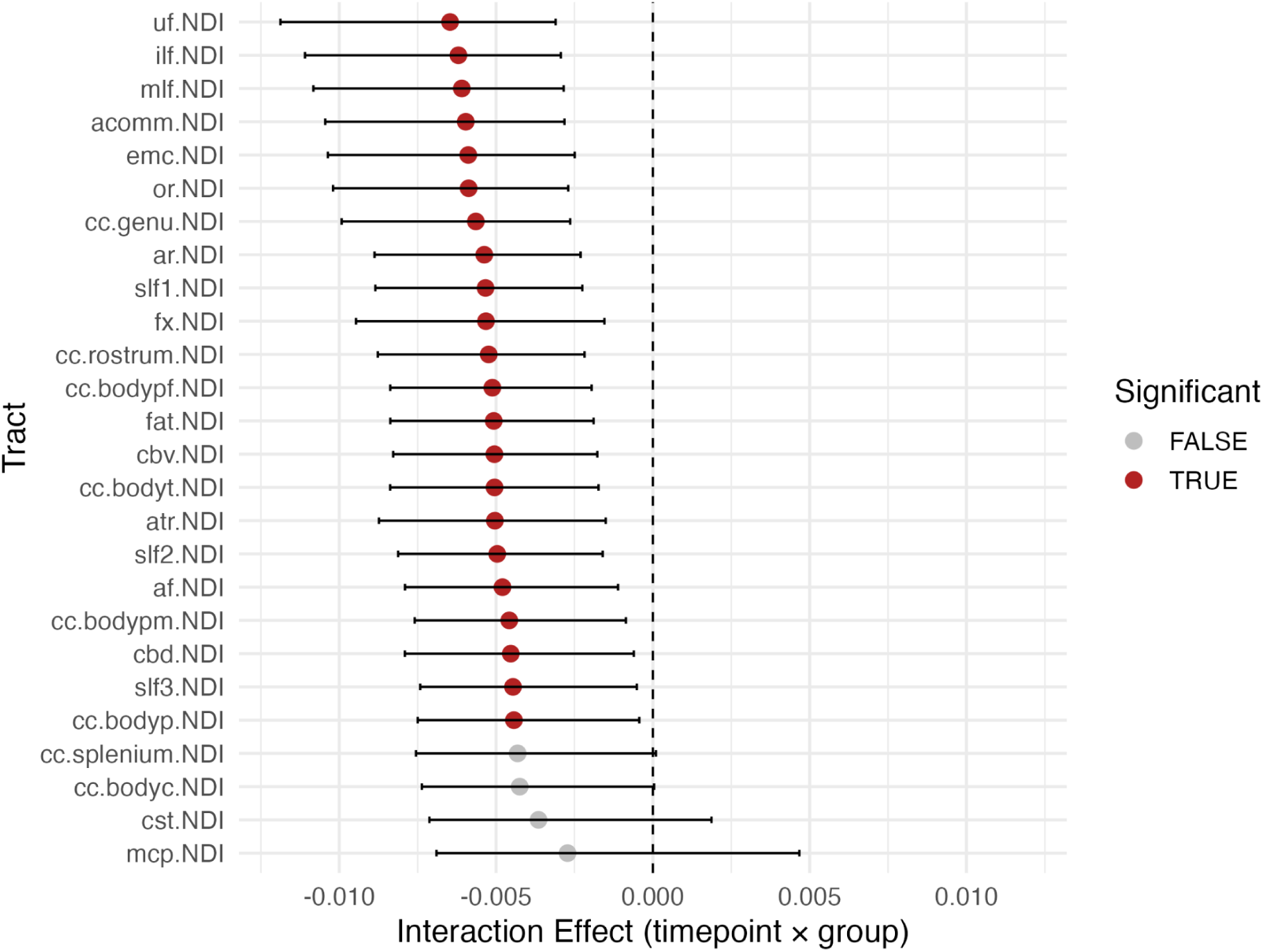
Estimates and 95% credible intervals of the Timepoint × Group interaction for Neurite Density Index (NDI) values across the 26 major white-matter tracts. Tracts for which the 95% credible interval does not include zero are considered to show evidence for an interaction effect. *Abbreviations*: uf: uncinate fasciculus; ilf: inferior longitudinal fasciculus; mlf: middle longitudinal fasciculus; acomm: anterior commissure; emc: extreme capsule; or: optic radiation; cc.genu: corpus callosum genu; ar: acoustic radiation; slf1: superior longitudinal fasciculus I; fx: fornix; cc.rostrum: corpus callosum rostrum; cc.bodypf: body prefrontal; fat: frontal aslant tract; cbv: cingulum bundle ventral; cc.bodyt: corpus callosum body temporal; atr: anterior thalamic radiation; slf2: superior longitudinal fasciculus II; af: arcuate fasciculus; cc.bodypm: corpus callosum premotor; cbd: cingulum bundle dorsal; slf3: superior longitudinal fasciculus III; cc.bodyp: corpus callosum body parietal; cc.splenium: corpus callosum splenium; cc.bodyc: corpus callosum central; cst: corticospinal tract; mcp: middle cerebellar peduncle.

For ODI (Figure 4), the Bayesian multilevel model did not provide evidence for the Timepoint × Group interaction (BF = 0.23; estimate: 0.000, 95% credible interval [-0.003, 0.004], pd = 58.09%; see Table 4-1).

**Figure 4.**
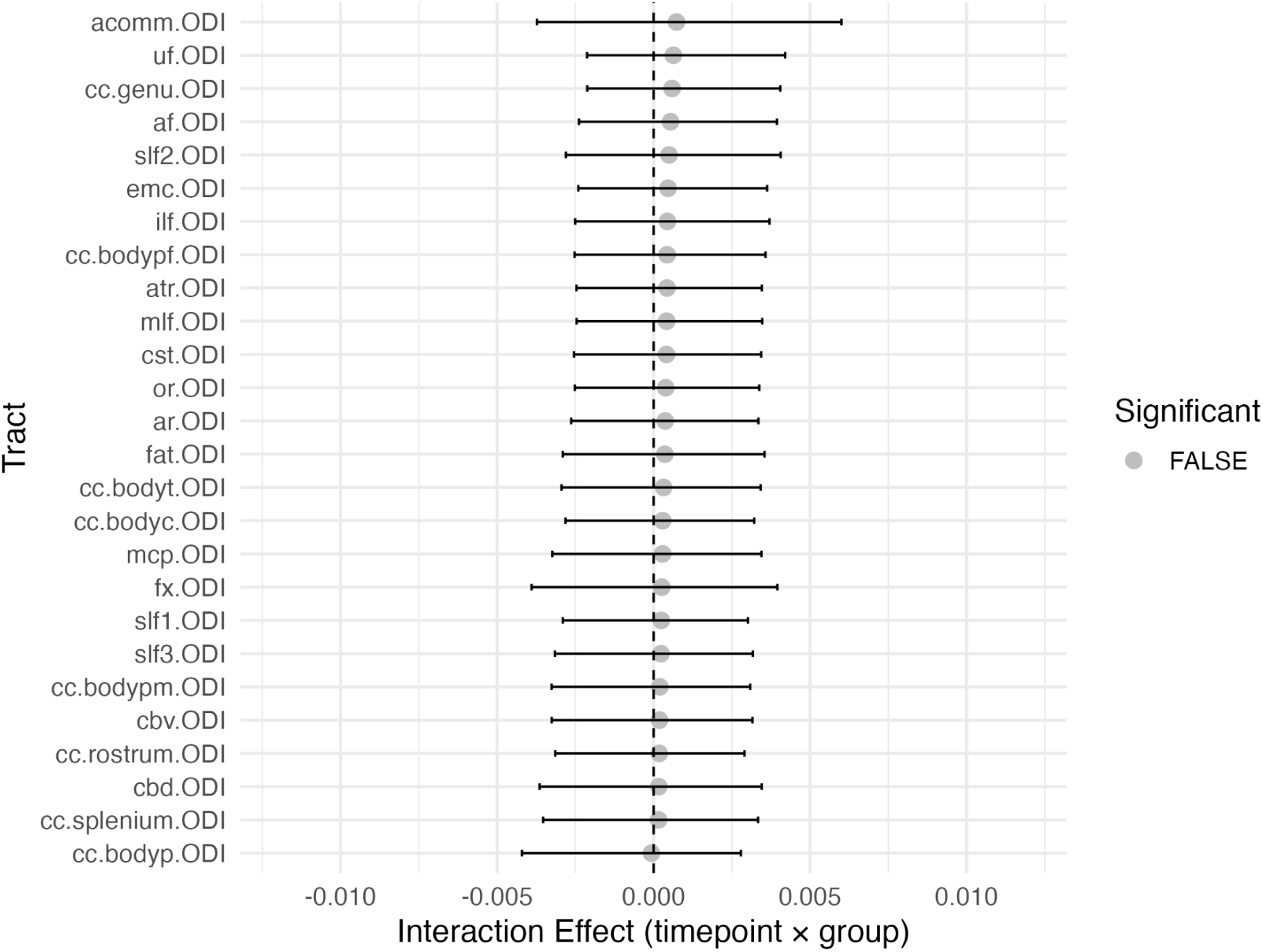
Estimates and 95% credible intervals of the Timepoint × Group interaction for Orientation Dispersion Index (ODI) across the 26 major white-matter tracts. For all tracts, the 95% credible interval includes zero, indicating no evidence for an interaction effect. *Abbreviations*: cbv: cingulum bundle ventral; cc.splenium: corpus callosum splenium; fx: fornix; cc.bodyt: cc: corpus callosum body temporal; cc.rostrum: corpus callosum rostrum; acomm: anterior commissure; uf: uncinate fasciculus; cc.bodyc: corpus callosum central; cc.genu: corpus callosum genu; atr: anterior thalamic radiation; slf1: superior longitudinal fasciculus I; mlf: middle longitudinal fasciculus; cc.bodypm: corpus callosum premotor; cc.bodyp: corpus callosum body parietal; slf3: superior longitudinal fasciculus III; cst: corticospinal tract; or: optic radiation; af: arcuate fasciculus; ar: acoustic radiation; ilf: inferior longitudinal fasciculus; slf2: superior longitudinal fasciculus II; fat: frontal aslant tract; mcp: middle cerebellar peduncle; emc: extreme capsule; cc.bodypf: corpus callosum prefrontal; cbd: cingulum bundle dorsal.

### 3.5. Relationship between changes in cardiorespiratory fitness and NODDI indices averaged across tracts

Change in NDI, averaged across all 26 major white-matter tracts, was positively associated with change in VO_2_max in all participants (r_(62)_ = 0.26, p = 0.037, 95% CI [0.02, 0.48]; also see Figure 3-1). However, the strength of this association did not differ significantly between the intervention groups (Fisher’s z = 0.31, p = 0.753).

Unlike for NDI, changes in ODI, averaged across all 26 major white matter tracts, were not significantly associated with changes in VO_2_max (r_(62)_ = 0.10, p = 0.419, 95% CI [-0.15, 0.34]; also see Figure 4-1). This association did not differ between groups (Fisher’s z = 0.73, p = 0.464).

### 3.6. Relationship between changes in NODDI indices averaged across tracts and cognition

Intervention-related improvements in inhibition and visual memory had previously been reported in the full trial (Schrenk et al., 2026). In the MRI subsample included in the present study, the intervention effect on inhibition remained significant, whereas the Timepoint × Group interaction for visual memory was not statistically significant (see Tables 1-2 and 1-3). Changes in neither inhibition nor visual memory were significantly associated with changes in NDI or ODI averaged across all 26 white matter tracts, either across all participants or specifically within the PAG (all uncorrected *p*-values > 0.250; Table 1-4).

### 3.7. Sensitivity Analyses

Head motion (tmi) did not systematically differ between or across (F_(1, 64)_ = 0.01, p = 0.914) groups (F_(1, 62)_ = 0.87, p = 0.355) and timepoints (F_(1, 64)_ = 0.25, p = 0.616). In line with that, both NDI and ODI model results remained the same when tmi was included as an additional covariate (NDI, Timepoint × Group: b = -0.005, SE = 0.002, t_(3338)_ = -2.93, p = 0.003; ODI, Timepoint × Group: b = -0.0005, SE = 0.002, t_(3338)_ = -0.27, p = 0.785). Finally, results also remained the same when a random term to account for pre-post correlations of the same tract in the same participant was added into the linear mixed effect model (NDI, Timepoint × Group: b = -0.005, SE = 0.001, t_(1714)_ = -5.47, p < 0.0001; ODI, Timepoint × Group: b = -0.0005, SE = 0.0005, t_(1714)_ = -1.14, p = 0.253).

## 4. Discussion

The present study provides evidence that an eight-week multicomponent physical exercise intervention is associated with increases in NODDI-derived neurite density in older adults. We found a significant Timepoint × Group interaction for NDI, reflecting a greater pre-to-post increase in the PAG than in the CON. Bayesian multilevel analyses further suggested that this intervention effect was broadly distributed across major white-matter tracts rather than being confined to a small number of isolated pathways. In contrast, no intervention effects were observed for ODI. Across all participants, increases in mean NDI were weakly associated with increases in cardiorespiratory fitness, whereas no consistent associations were observed between changes in NODDI indices and cognitive performance. Together, these results indicate that a relatively short multicomponent physical exercise intervention may induce selective changes in white matter microstructure, as reflected by increases in neurite density. By demonstrating intervention-related changes in NDI, the present study extends the neuroimaging findings from the FIT4BRAIN RCT (Saraei et al., 2025; Schrenk et al., 2026) to a complementary diffusion-MRI-derived marker of white-matter microstructure.

In the Bayesian multilevel model, the tract-specific posterior estimates of the Timepoint × Group interaction had 95% credible intervals excluding zero for 22 of the 26 tracts. This widespread pattern suggests that the exercise-related increases in NDI were not confined to specific pathways but occurred across much of the white matter. Rather than indicating localized plasticity, these findings are consistent with the possibility that multicomponent physical exercise promotes more distributed microstructural adaptations. Similar widespread effects of physical exercise on white matter have been previously suggested based on diffusion tensor imaging metrics (Voss et al., 2013). The present findings extend this literature by showing that comparable intervention-related changes can also be detected using the NODDI-derived neurite density index.

The intervention-related effects were specific to NDI, whereas no corresponding effects were observed for ODI. This dissociation suggests that the intervention preferentially affected aspects of the diffusion signal captured by NDI rather than the spatial dispersion of neurite organization. The absence of ODI changes therefore indicates that the observed microstructural adaptations were selective rather than affecting all NODDI-derived indices. One possible interpretation is that neurite density and orientation dispersion capture distinct aspects of white-matter microstructure that may differ in their responsiveness to physical exercise.It is also possible that methodological factors, including the relatively narrow range of ODI values in the present homogeneous sample, may have limited the detection of small changes. Future studies including participants with a broader range of ages, cardiorespiratory fitness levels, and health statuses may help clarify whether physical exercise-related changes in orientation dispersion emerge in more heterogeneous populations.

Our second hypothesis regarding the relationship between changes in NODDI indices and cardiorespiratory fitness was only partially supported. Across participants, changes in mean NDI, but not mean ODI, were positively correlated with changes in cardiorespiratory fitness. However, the strength of this association did not differ significantly between the PAG and CON. Thus, the pooled association cannot be interpreted as intervention-specific. Nevertheless, it is broadly consistent with previous evidence linking cardiorespiratory fitness to white matter integrity (Voss et al., 2013; Mendez Colmenares et al., 2021; Polk et al., 2023).

The absence of a statistically significant difference between the group-specific correlations, together with the non-significant Timepoint × Group interaction for VO_2_max, limits conclusions about cardiorespiratory fitness as a mechanism underlying the observed intervention effect on NDI. Although VO_2_max increased from pre- to post-intervention within the PAG, the change was not significantly greater than that observed in the CON. Therefore, the present findings cannot establish or exclude changes in cardiorespiratory fitness as a mediator of the intervention-related NDI changes. The relatively high baseline fitness of the sample may have limited the potential for further improvement. In addition, the intensity or volume of the intervention, which was designed to be appropriate for older adults, may not have provided a sufficient training stimulus to elicit substantial cardiorespiratory adaptations in comparatively fit participants. However, these interpretations remain speculative. Although physical exercise may influence white matter microstructure through mechanisms associated with improvements in cardiorespiratory fitness, including enhanced vascular health and cerebral perfusion (Sexton et al., 2016), it may also exert more direct effects through exercise-induced peripheral signaling molecules linked to neuroplasticity and angiogenesis (Tari et al., 2025). Consequently, improvements in white matter microstructure may not depend exclusively on measurable changes in cardiorespiratory fitness (Burzynska et al., 2014). However, the present study was not specifically designed to distinguish among these potential mechanisms.

We found no evidence that changes in NDI or ODI were associated with changes in inhibition performance or visual memory. Although an intervention-related improvement in inhibition was also observed in the present MRI subsample, individual changes in inhibition did not covary with individual changes in NODDI indices. The smaller size of the MRI subsample compared with the full randomized trial likely reduced the power both to reproduce all previously observed cognitive intervention effects and, particularly, to detect associations between neuroimaging and cognitive change scores. Thus, the absence of significant brain–behavior associations should not be interpreted as evidence that exercise-related white matter changes are unrelated to cognitive outcomes. Rather, it is consistent with the heterogeneous evidence regarding relationships between exercise-related brain changes and cognitive outcomes (Di Lorito et al., 2021; Silva et al., 2024). Longer interventions or follow-up periods may be required before relations between changes in white matter microstructure and cognition are detectable (Mendez Colmenares et al., 2021; Polk et al., 2023; Chen et al., 2025). Such associations may also be more pronounced in populations with greater cognitive variability or increased risk for cognitive decline, such as individuals with subjective cognitive decline or mild cognitive impairment (Whitfield et al., 2021; Kušleikienė et al., 2025).

The results of the present study should be considered in light of several limitations. First, the present NODDI analyses focused on secondary neuroimaging outcomes, and the study was not specifically powered to detect changes in NDI or ODI. Second, NDI is a model-derived diffusion metric and should not be interpreted as a direct histological measure of dendritic or axonal density. Third, the predominance of females (>60%) and the relatively high cardiorespiratory fitness level of our participants at baseline may limit the generalizability of our findings while precluding a meaningful examination of potential sex differences in outcomes. Finally, longer-term assessments are needed to determine whether the observed NDI changes persist after the intervention. Despite these limitations, the randomized design, active control condition, longitudinal diffusion imaging, and converging frequentist and Bayesian analyses provide a robust foundation for further investigations of exercise-related changes in NODDI-derived white matter indices. Future studies should investigate the reproducibility of these findings in larger and more diverse samples and across different intervention durations, fitness levels, and cognitive risk profiles.

To conclude, eight weeks of a multicomponent physical exercise intervention resulted in greater increases in neurite density across most major white matter tracts in healthy older adults compared with an active control intervention. In contrast, no comparable effects were observed for orientation dispersion. Together, these findings suggest that NDI may provide a useful marker of exercise-related white matter plasticity in older adults. Future studies should determine whether the observed microstructural changes persist over time and whether they translate into measurable cognitive improvements following longer interventions or in populations at increased risk of cognitive decline.

## Data Availability

The data for the main analyses can be accessed before manuscript publication at peer-review-link.

https://zenodo.org/records/22206492?preview=1&token=eyJhbGciOiJIUzUxMiIsImlhdCI6MTc4ODE3NjI1NCwiZXhwIjoxODAxMzUzNTk5fQ.eyJpZCI6ImU0NjBmYTdlLTNmNjctNDg5ZC1hZDQxLWZjMGE5YTg1MjViNSIsImRhdGEiOnt9LCJyYW5kb20iOiI4ZGMxNjE0MDlhMGM5YjhkOGJkZTBmNmM4YTY4YTc4NyJ9.rCV0JYpHp8sjB8WydrrdHm5robm6pz_mZQNdtnQtEKeSs7-s6PwbFmzaD3R54QHooWlohmJ76XYourjAenkUVQ

## Author contributions

Conceptualization: A.R.R., S.B., K.F.

Data Curation: A.R.R., S.B., S.J.S.

Formal Analysis: A.R.R.

Funding Acquisition: C.F., C.G., O.W.W., K.F.

Investigation: S.J.S., M.H.

Methodology: A.R.R., K.F.

Project Administration: C.F.

Resources: C.P.

Software: A.R.R.

Supervision: K.F.

Validation: A.R.R., S.J.S., K.F.

Visualization: A.R.R.

Writing – original draft: A.R.R.

Writing – review & editing: A.R.R., S.J.S., M.H., C.P., K.F.

## Conflicts of Interest

The authors declare no conflicts of interest

## Acknowledgments

This study was funded by the European Union’s Horizon 2020 research and innovation program under the Marie Skłodowska-Curie grant agreements No. 859890 (SmartAge), No. 101227102 (MenoBrain), the German Research Foundation (DFG; FI 1424/2-2), and the IZKF Advanced Medical Scientist Program of Jena University Hospital. The authors wish to thank the high-performance computing cluster at Friedrich-Schiller-University Jena for providing the resources to efficiently conduct the MRI analyses for this study.

## Notes

### Competing Interest Statement

The authors have declared no competing interest.

### Clinical Trial

DRKS00028022

### Author Declarations

The ethics committee of Jena University Hospital gave ethical approval for this work (No. 2021-2345-BO).

