## Extended data for "Physical exercise increases the NODDI-derived neurite density index across white matter tracts in healthy older adults: results from the FIT4BRAIN randomized controlled trial"

### Methods

For the Bayesian models, results were deemed ‘significant’ if the Bayes factor (BF) ≥ 10 or the posterior probability of direction (pd) ≥ 95%.

### Results

#### Sex differences and correlations with age at baseline

At baseline, independent of intervention group, females (versus males) had significantly higher mean $VO_2$ max z-scores (1.68 ± 0.95 vs. 0.71 ± 0.48, respectively), lower mean NDI (-0.21 ± 0.75 vs. 0.42 ± 0.66, respectively), lower waist-to-hip ratio (0.81 ± 0.07 vs. 0.96 ± 0.08, respectively), lower total intracranial volume (1,487 ± 110 vs. 1,682 ± 127 $cm^3$, respectively), lower white matter volume (409.84 ± 44.0 vs. 472.48 ± 46.87, respectively), lower hippocampal volume (3.94 ± 0.40 vs. 4.29 ± 0.33), and greater head motion during DWI (0.59 ± 1.37 vs. -0.41 ± 0.96, respectively; all p-values < 0.001). Mean NDI, but not mean ODI ($r_{66}$ = -0.03, p = 0.826), significantly correlated with age, with older age associated with lower mean NDI values ($r_{66}$ = -0.32, p = 0.009). Neither mean NDI ($r_{65}$ = -0.04, p = 0.737) nor mean ODI ($r_{65}$ = -0.004, p = 0.970) correlated with $VO_2$max at baseline. No remaining significant sex differences (all p-values > 0.4) or correlations with age (all p-values > 0.075) were observed.

#### Mean heart rate (average and maximum) achieved during intervention

Table 1-1 shows the results of an analysis of covariance (ANCOVA) investigating the effects of intervention (i.e., physical exercise/activity: PAG or control intervention: CON) on (a) the average and (b) the maximum heart rate across conducted units, while controlling for the effects of age and sex.

**Table 1-1.** ANCOVA model results for the average and maximum heart rate (beats per minute) across intervention units per group while controlling for age and sex

| **Variable** | **PAG**[a] **(*n* = 33)** | **CON**[b] **(*n* = 27)** | ***F*-value (*df*1, *df*2)** | ***p*-value** |
|---|---|---|---|---|
| *Mean heart rate* | | | | |
| Walking (PAG) vs. progressive muscle relaxation (CON) | 106.46 ± 8.95 | 69.74 ± 8.36 | 259.14 (1, 56) | **< 0.0001** |
| Age | - | - | 0.46 (1, 56) | 0.498 |
| Sex | - | - | 0.26 (1, 56) | 0.611 |
| Yoga (PAG) vs. progressive muscle relaxation (CON) | 77.65 ± 7.26 | 69.74 ± 8.36 | 15.31 (1, 55) | **0.0002** |
| Age | - | - | 0.40 (1, 55) | 0.531 |
| Sex | - | - | 2.31 (1, 55) | 0.135 |
| *Maximum heart rate* | | | | |
| Walking (PAG) vs. progressive muscle relaxation (CON) | 129.50 ± 9.49 | 85.51 ± 9.57 | 309.49 (1, 56) | **< 0.0001** |
| Age | - | - | 0.01 (1, 56) | 0.900 |
| Sex | - | - | 0.60 (1, 56) | 0.439 |
| Yoga (PAG) vs. progressive muscle relaxation (CON) | 90.65 ± 8.03 | 85.51 ± 9.57 | 5.06 (1, 55) | **0.028** |
| Age | - | - | 0.000001 (1, 55) | 0.999 |
| Sex | - | - | 2.88 (1, 55) | 0.095 |

*Note*: [a] One and [b] five missing data points due to unrecorded data equipment failure (e.g., the watch broke and was replaced, but syncing did not work or the participant could not figure out how to use the watch).

Effect of intervention type on neurite density and orientation dispersion on individual major white-matter tracts

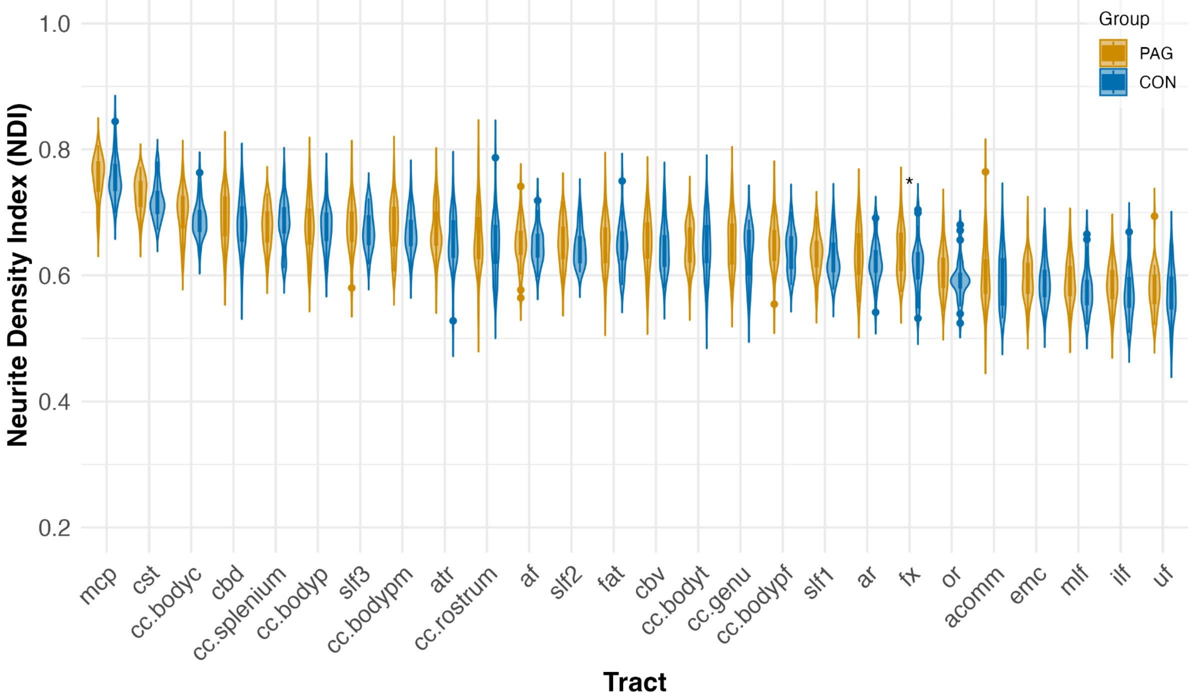


**Figure 1-1.** Neurite Density Index (NDI) values for both groups at baseline. Violin plots show the distribution of NDI values for the physical exercise intervention ("PAG", in dark yellow) and the active control group ("CON", in blue). Dots represent outlier observations. Tracts are ordered by descending NDI values across participants. * Asterisks indicate nominally significant between-group differences ($p_{uncorrected} < 0.05$), which did not remain significant after FDR correction. *Abbreviations*: mcp: middle cerebellar peduncle; cst: corticospinal tract; cc.bodyc: corpus callosum central; cbd: cingulum bundle dorsal; cc.splenium/.bodyp: corpus callosum splenium/body parietal; slf3: superior longitudinal fasciculus III; cc.bodypm: corpus callosum premotor; atr: anterior thalamic radiation; cc.rostrum: corpus callosum rostrum; af: arcuate fasciculus; slf2: superior longitudinal fasciculus II; fat: frontal aslant tract; cbv: cingulum bundle ventral; cc.bodyt/genu/bodypf: corpus callosum body temporal/genu/body prefrontal; slf1: superior longitudinal fasciculus I; ar: acoustic radiation; fx: fornix; or: optic radiation; acomm: anterior commissure; emc: extreme capsule; mlf: middle longitudinal fasciculus; ilf: inferior longitudinal fasciculus; uf: uncinate fasciculus.

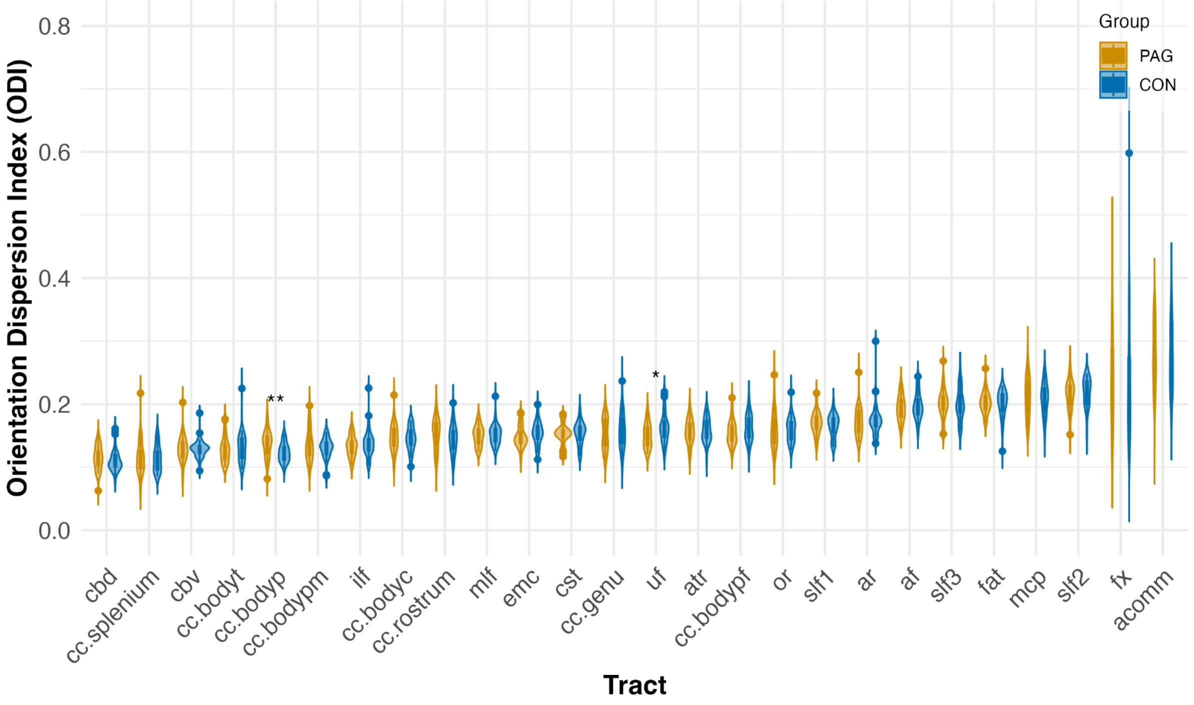


**Figure 1-2**. Orientation Dispersion Index (ODI) values for both groups at baseline. Violin plots of ODI distributions are shown for the physical exercise intervention ("PAG", in dark yellow) and the active control intervention ("CON", in blue). Dots represent outlier observations. Tracts are ordered by ascending ODI values across participants. * Asterisks indicate nominally significant between-group differences ($p_{uncorrected}$ < 0.05, ** $p_{uncorrected}$ < 0.01), none of which remained significant after FDR correction. *Abbreviations*: cbd: cingulum bundle dorsal; cc.splenium: corpus callosum splenium; cbv: cingulum bundle ventral; cc.bodyt/bodyp/bodypm: corpus callosum body temporal/body parietal/body premotor; ilf: inferior longitudinal fasciculus; cc.bodyc: corpus callosum central; cc.rostrum: corpus callosum rostrum; mlf: middle longitudinal fasciculus; emc: extreme capsule; cst: corticospinal tract; cc.genu: corpus callosum genu; uf: uncinate fasciculus; atr: anterior thalamic radiation; cc.bodypf: corpus callosum body prefrontal; or: optic radiation; slf1: superior longitudinal fasciculus I; ar: acoustic radiation; af: arcuate fasciculus; slf3: superior longitudinal fasciculus III; fat: frontal aslant tract; mcp: middle cerebellar peduncle; slf2: superior longitudinal fasciculus II; fx: fornix;  acomm: anterior commissure.

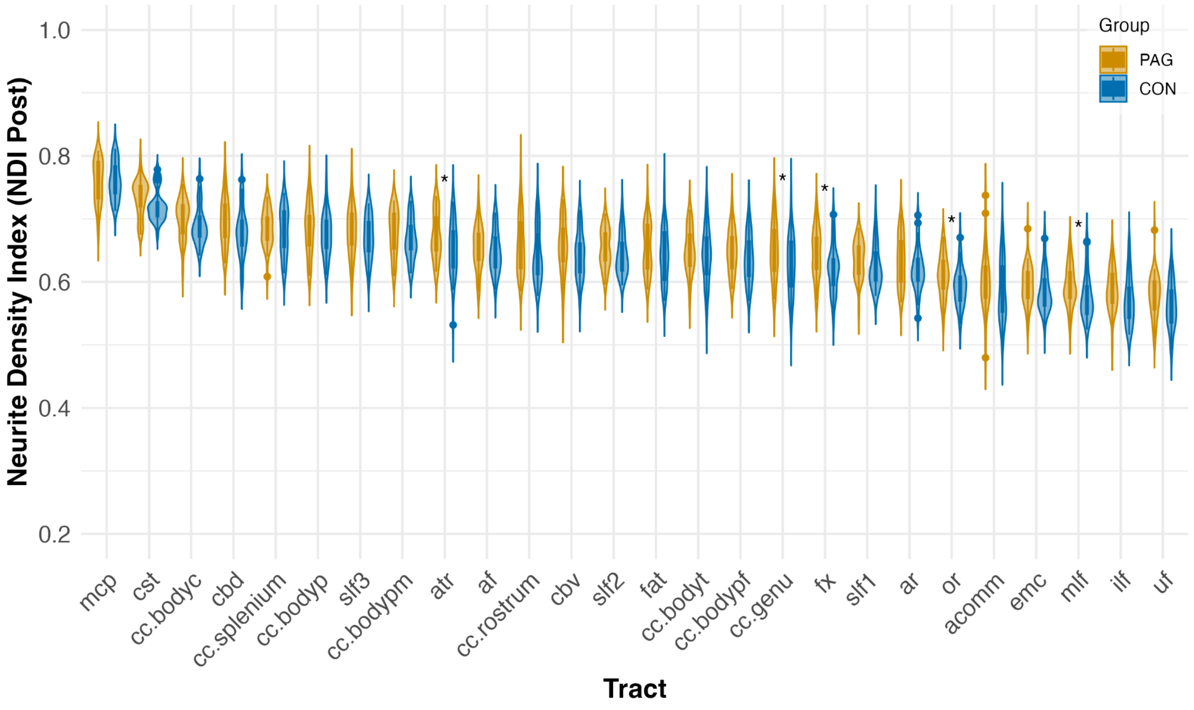


**Figure 1-3.** Mean Neurite Density Index values post-intervention ("NDI Post") for both groups across 26 major white matter tracts. The physical exercise intervention ("PAG") is depicted in dark yellow, whereas the control intervention ("CON") is shown in blue. Tracts are ordered by descending NDI value. * Asterisks indicate significant group differences ($p_{uncorrected} < 0.05$). *Abbreviations*: mcp: middle cerebellar peduncle; cst: corticospinal tract; cc.bodyc: corpus callosum central; cbd: cingulum bundle dorsal; cc.splenium/.bodyp: corpus callosum splenium/body parietal; slf3: superior longitudinal fasciculus III; cc.bodypm: corpus callosum premotor; atr: anterior thalamic radiation; af: arcuate fasciculus; cc.rostrum: corpus callosum rostrum; cbv: cingulum bundle ventral; slf2: superior longitudinal fasciculus II; fat: frontal aslant tract; cc.bodyt/bodypf/genu: corpus callosum body temporal/body prefrontal/genu; fx: fornix; slf1: superior longitudinal fasciculus I; ar: acoustic radiation; or: optic radiation; acomm: anterior commissure; emc: extreme capsule; mlf: middle longitudinal fasciculus; ilf: inferior longitudinal fasciculus; uf: uncinate fasciculus.

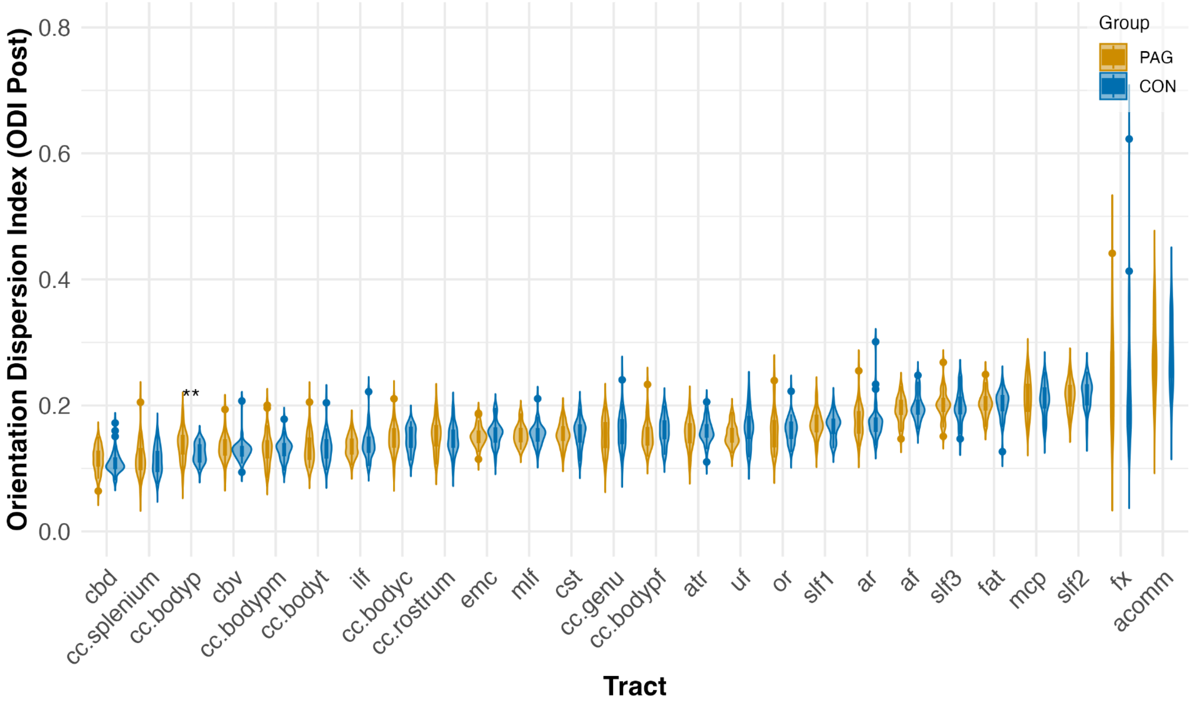


**Figure 1-4.** Mean Orientation Dispersion Index values post-intervention ("ODI Post") for both groups across 26 major white matter tracts. The physical exercise intervention ("PAG") is depicted in dark yellow, whereas the control intervention ("CON") is shown in blue. Tracts are ordered by ascending ODI values. ** $p_{uncorrected} < 0.01$. *Abbreviations*: cbd: cingulum bundle dorsal; cc.splenium: corpus callosum splenium; cc.bodyp: corpus callosum body parietal; cbv: cingulum bundle ventral; cc.bodypm/bodyt: corpus callosum body premotor/body temporal; ilf: inferior longitudinal fasciculus; cc.bodyc: corpus callosum central; cc.rostrum: corpus callosum rostrum; emc: extreme capsule; mlf: middle longitudinal fasciculus; cst: corticospinal tract; cc.genu: corpus callosum genu; cc.bodypf: corpus callosum body prefrontal; atr: anterior thalamic radiation; uf: uncinate fasciculus; or: optic radiation; slf1: superior longitudinal fasciculus I; ar: acoustic radiation; af: arcuate fasciculus; slf3: superior longitudinal fasciculus III; fat: frontal aslant tract; mcp: middle cerebellar peduncle; slf2: superior longitudinal fasciculus II; fx: fornix; acomm: anterior commissure.

### Correlation between Neurite Density Index (NDI) and Orientation Dispersion Index (ODI)

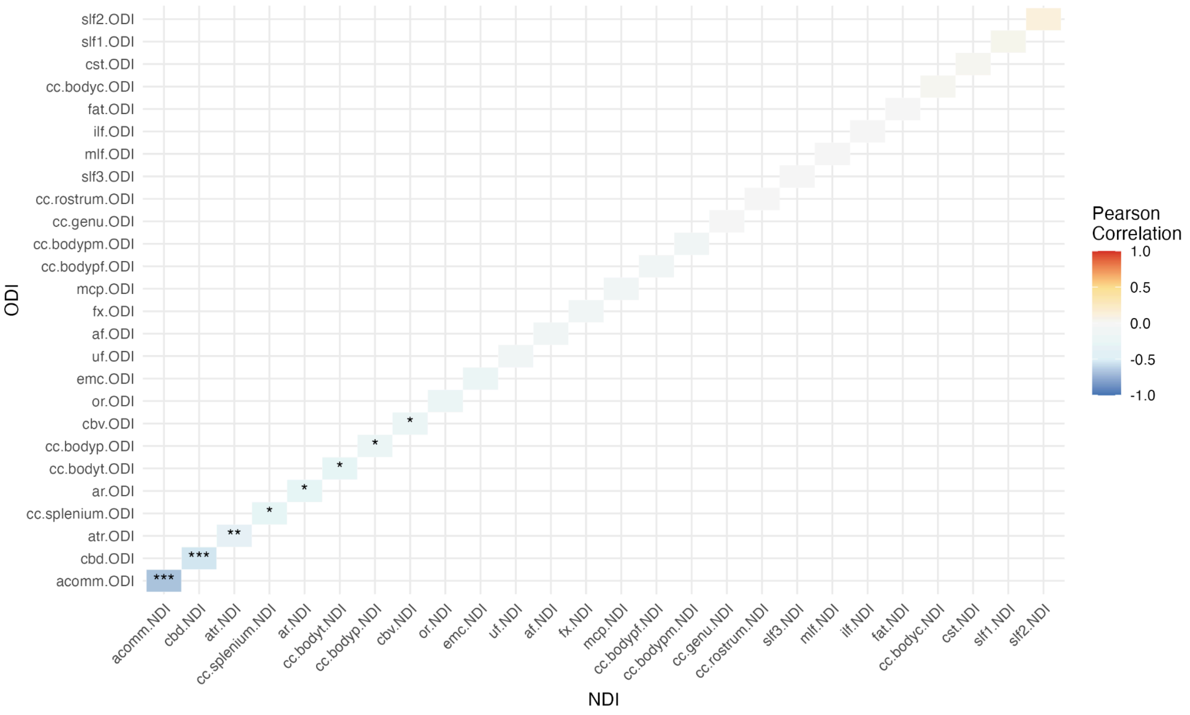


**Figure 1-5**. Correlation between ODI and NDI values at baseline. * p < 0.05, ** p < 0.01, *** p < 0.001

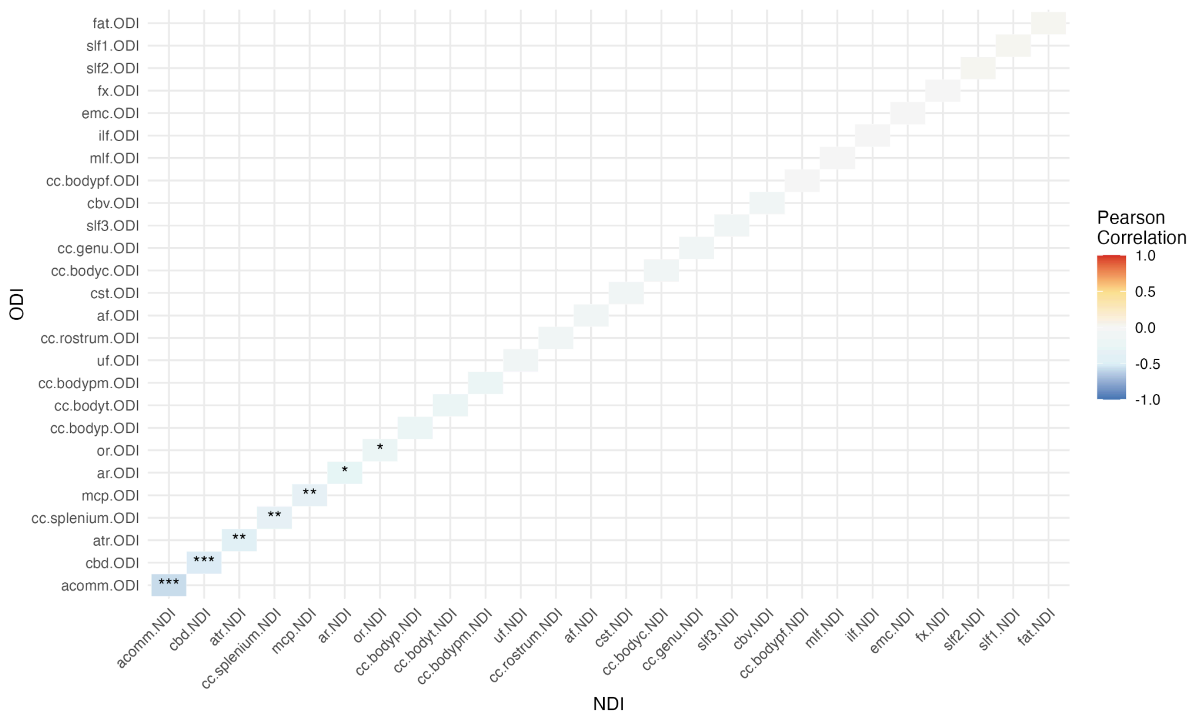


**Figure 1-6**. Correlation between ODI and NDI values post-intervention. * $p < 0.05$, ** $p < 0.01$, *** $p < 0.001$

### Bayesian multilevel model results

**Table 3-1.** Results of the Bayesian multilevel model on the neurite density index (NDI)

| Fixed Effects | Estimate | 95% Credible Interval | pd | BF |
|---|---|---|---|---|
| Timepoint (Post) | 0.003 | [0.001, 0.005] | **99.32%** | **1.56** |
| Group (CON) | -0.01 | [-0.02, 0.01] | 83.69% | 0.46 |
| Sex (Males) | 0.02 | [0.01, 0.03] | **99.99%** | **68.5** |
| Age | -0.002 | [-0.004, -0.001] | **99.83%** | **5.75** |
| Timepoint × Group | -0.005 | [-0.01, -0.002] | **99.88%** | **21.11** |

*Note*. Model: NDI ~ timepoint * group + sex + age + (1 + timepoint * group | tract) + (1 | id)

The Bayes Factor (BF) indicates the data-based strength of evidence for each of the fixed effects on NDI: BF < 1: 'Evidence for null'; BF < 3: 'Anecdotal'; BF < 10: 'Moderate'; BF < 30: 'Strong'; BF < 100: 'Very strong'. The credible interval indicates that there is a 95% probability that the true parameter value lies within this interval, given the current model and priors. The posterior probability of direction (pd) indicates the proportion of the posterior distribution that falls on the same side of zero as the median. Values pd ≥ 95% or BF ≥ 10 are marked in boldface to indicate 'significance.'

**Table 4-1.** Results of the Bayesian multilevel model on the orientation dispersion index (ODI)

| Fixed Effects | Estimate | 95% Credible Interval | pd | BF |
|---|---|---|---|---|
| Timepoint (Post) | 0.001 | [-0.002, 0.003] | 68.67% | 0.09 |
| Group (CON) | 0.000 | [-0.004, 0.004] | 55.18% | 0.04 |
| Sex (Males) | 0.000 | [-0.004, 0.003] | 56.39% | 0.03 |
| Age | 0.000 | [-0.001, 0] | 69.59% | 0.03 |
| Timepoint × Group | 0.000 | [-0.003, 0.004] | 58.09% | 0.23 |

*Note*. Model: ODI ~ timepoint * group + sex + age + (1 + timepoint * group | tract) + (1 | id)

The Bayes Factor (BF) indicates the data-based strength of evidence for each of the fixed effects on NDI: BF < 1: 'Evidence for null'; BF < 3: 'Anecdotal'; BF < 10: 'Moderate'; BF < 30: 'Strong'; BF < 100: 'Very strong'. The credible interval indicates that there is a 95% probability that the true parameter value lies within this interval, given the current model and priors. The posterior probability of direction (pd) indicates the proportion of the posterior distribution that falls on the same side of zero as the median.

### Relationship between changes in cardiorespiratory fitness and NODDI indices across tracts

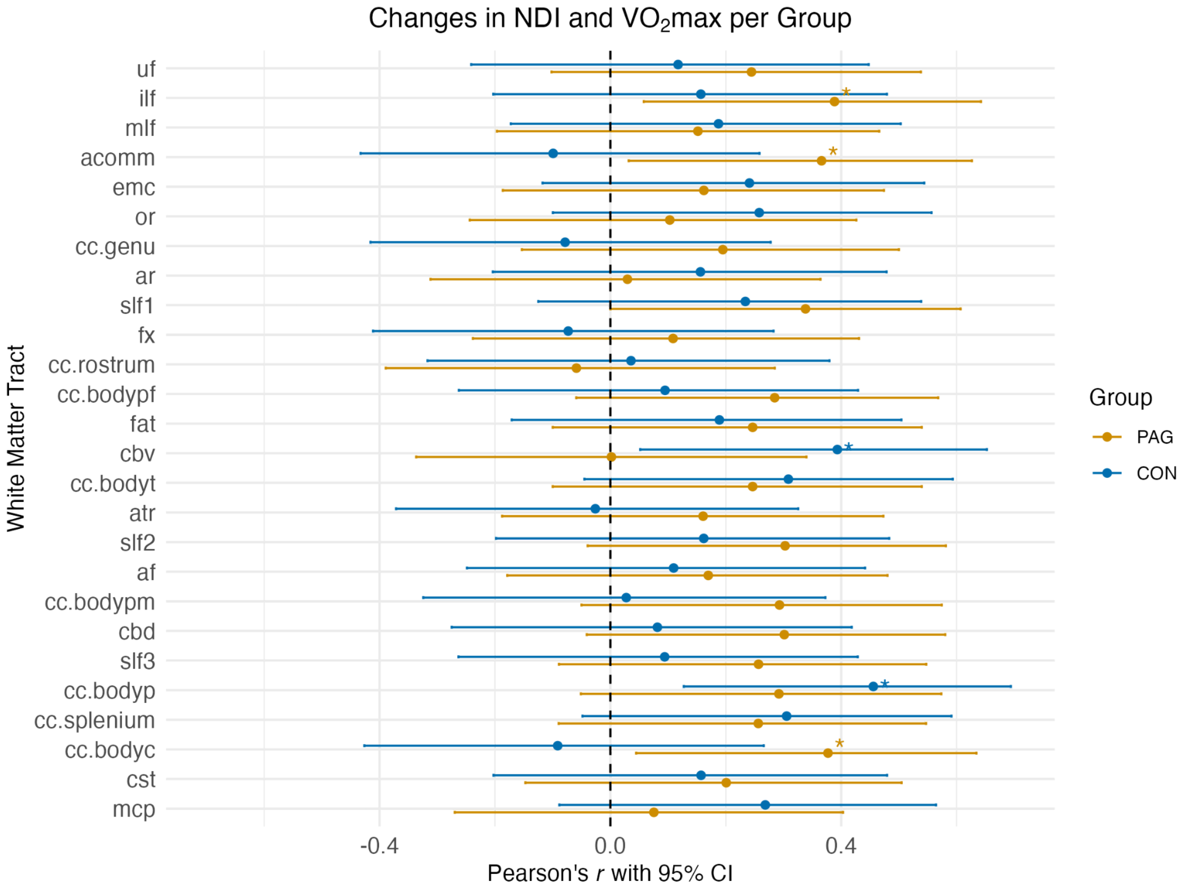


**Figure 3-1.** Pearson's correlation coefficient (r) between changes in Neurite Density Index (NDI) and changes in cardiorespiratory fitness (mean $VO_2max$) across 26 major white matter tracts. Changes represent the post-pre intervention differences and are shown separately for the physical activity intervention ("PAG"; in dark yellow) and control ("CON"; in blue) groups. Dots indicate correlation coefficients, and the horizontal lines represent 95% confidence intervals (CI). Tracts are ordered as in Figure 3. * Asterisks indicate correlations at $p_{uncorrected} < 0.05$. *Abbreviations*: uf: uncinate fasciculus; ilf: inferior longitudinal fasciculus; mlf: middle longitudinal fasciculus; acomm: anterior commissure; emc: extreme capsule; or: optic radiation; cc.genu: corpus callosum genu; ar: acoustic radiation; slf1: superior longitudinal fasciculus I; fx: fornix; cc.rostrum: corpus callosum rostrum; cc.bodypf: body prefrontal; fat:

frontal aslant tract; cbv: cingulum bundle ventral; cc.bodyt: corpus callosum body temporal; atr: anterior thalamic radiation; slf2: superior longitudinal fasciculus II; af: arcuate fasciculus; cc.bodypm: corpus callosum premotor; cbd: cingulum bundle dorsal; slf3: superior longitudinal fasciculus III; cc.bodyp: corpus callosum body parietal; cc.splenium: corpus callosum splenium; cc.bodyc: corpus callosum central; cst: corticospinal tract; mcp: middle cerebellar peduncle.

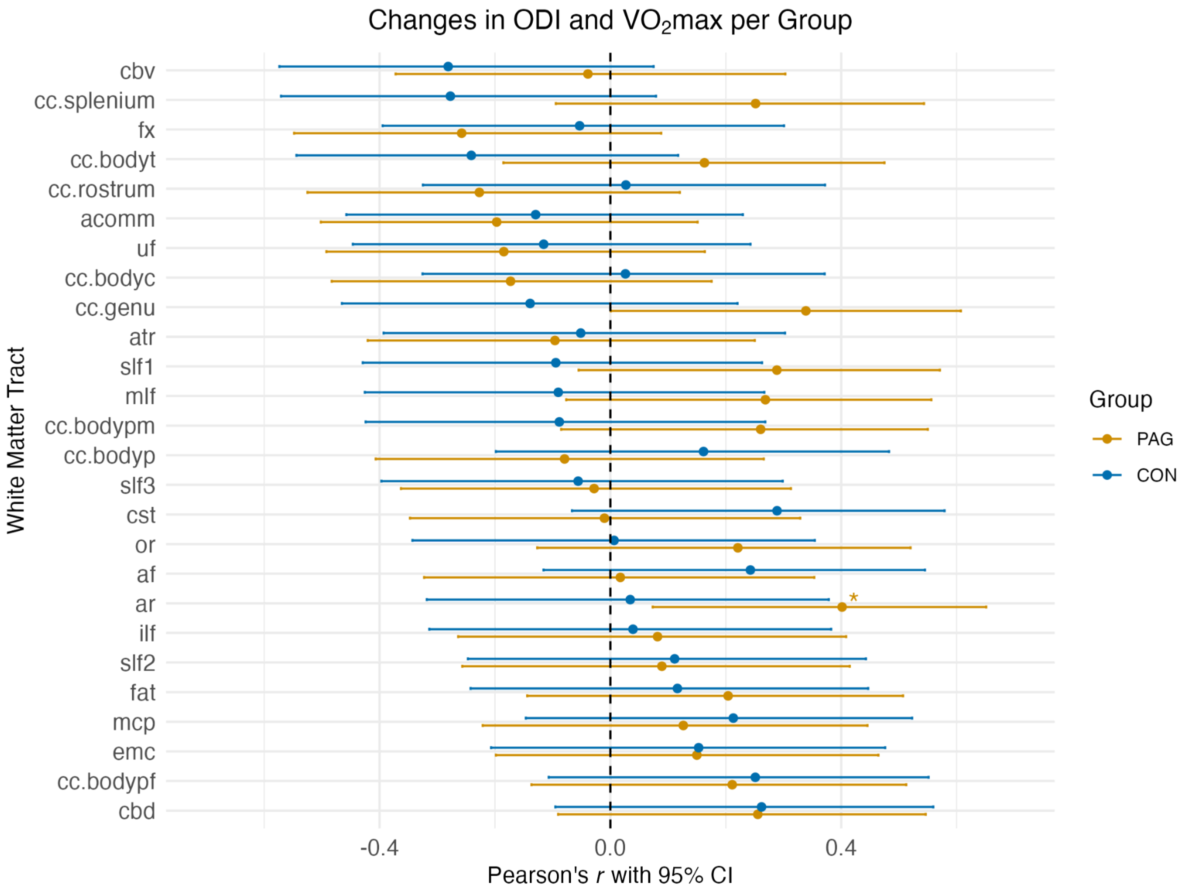


**Figure 4-1.** Pearson's correlation coefficient (r) between changes in Orientation Dispersion Index (ODI) and changes in cardiorespiratory fitness (mean $VO_2$ max) across 26 major white matter tracts. Changes represent the post-pre intervention differences and are shown separately for the physical activity intervention ("PAG"; in dark yellow) and control ("CON"; in blue) groups. Dots indicate correlation coefficients, and the lines represent 95% confidence intervals (CI). Tracts are ordered by ascending correlation coefficient. * Asterisks indicate significant group differences ($p_{uncorrected}$ < 0.05). *Abbreviations*: cbv: cingulum bundle ventral;

cc.splenium: corpus callosum splenium; fx: fornix; cc.bodyt: cc: corpus callosum body temporal; cc.rostrum: corpus callosum rostrum; acomm: anterior commissure; uf: uncinate fasciculus; cc.bodyc: corpus callosum central; cc.genu: corpus callosum genu; atr: anterior thalamic radiation; slf1: superior longitudinal fasciculus I; mlf: middle longitudinal fasciculus; cc.bodypm: corpus callosum premotor; cc.bodyp: corpus callosum body parietal; slf3: superior longitudinal fasciculus III; cst: corticospinal tract; or: optic radiation; af: arcuate fasciculus; ar: acoustic radiation; ilf: inferior longitudinal fasciculus; slf2: superior longitudinal fasciculus II; fat: frontal aslant tract; mcp: middle cerebellar peduncle; emc: extreme capsule; cc.bodypf: corpus callosum prefrontal; cbd: cingulum bundle dorsal.

### Effect of the intervention on cognitive variables in the current sample

**Table 1-2.** Results of the mixed-effects model on Stroop test inhibition score

| Fixed Effect | Estimate | SE | df | t-value | p-value | 95% CI |
|---|---|---|---|---|---|---|
| Timepoint (Post) | -8.31 | 3.12 | 63.66 | -2.66 | **0.010** | **[-14.42, -2.20]** |
| Group (CON) | -4.16 | 4.19 | 99.80 | -0.99 | 0.323 | [-12.36, 4.05] |
| Sex (Males) | 0.48 | 3.77 | 63.38 | 0.13 | 0.898 | [-6.90, 7.87] |
| Age | 0.85 | 0.50 | 64.65 | 1.69 | 0.096 | [-0.14, 1.84] |
| Timepoint × Group | 9.63 | 4.35 | 61.68 | 2.21 | **0.031** | **[1.10, 18.15]** |

*Note.* The reference levels for the first three variables are Pre, PAG, and females, from top to bottom. The timepoint estimate represents the pre-to-post change in the PAG group. Model: Stroop inhibition score ~ timepoint * group + sex + age + (1 | id). Significant fixed effects are highlighted in boldface. Degrees of freedom and p values for individual coefficients were calculated using the Satterthwaite approximation. CI: confidence interval; df: degrees of freedom; SE: Standard error.

**Table 1-3.** Results of the mixed-effects model on Rey-Osterrieth Figure memory score

| Fixed Effect | Estimate | SE | df | t-value | p-value | 95% CI |
|---|---|---|---|---|---|---|
| Timepoint (Post) | 17.62 | 1.91 | 64 | 9.20 | **<0.0001** | **[13.87, 21.37]** |
| Group (CON) | -2.74 | 2.64 | 102.9 | -1.04 | 0.302 | [-7.91, 2.43] |
| Sex (Males) | -1.75 | 2.36 | 62 | -0.74 | 0.461 | [-6.37, 2.87] |
| Age | 0.43 | 0.31 | 62 | 1.39 | 0.170 | [-0.18, 1.05] |
| Timepoint × Group | -3.48 | 2.75 | 64 | -1.26 | 0.210 | [-8.87, 1.91] |

*Note.* The reference levels for the first three variables are Pre, PAG, and females, from top to bottom. The timepoint estimate represents the pre-to-post change in the PAG group. Model: Rey-Osterrieth Figure memory score ~ timepoint * group + sex + age + (1 | id). Significant fixed effects are highlighted in boldface. Degrees of freedom and p values for individual coefficients were calculated using the Satterthwaite approximation. CI: confidence interval; df: degrees of freedom; SE: Standard error.

### Relationship between change in NDI/ODI and change in cognition due to intervention

**Table 1-4.** Bivariate correlations between changes in cognition and changes in NODDI indices across participants and separately for each intervention group

| Correlation | All | PAG | CON |
|---|---|---|---|
| NDI and FWIT Interference (changes) | r(59) = -0.04<br>p = 0.710<br>95% CI [-0.30, 0.20] | r(28) = 0.12<br>p = 0.543<br>95% CI [-0.26, 0.47] | r(31) = -0.07<br>p = 0.695<br>95% CI [-0.42, 0.29] |
| NDI and Rey-Osterrieth memory (changes) | r(66) = 0.12<br>p = 0.336<br>95% CI [-0.12, 0.35] | r(34) = 0.14<br>p = 0.424<br>95% CI [-0.21, 0.46] | r(32) = 0.03<br>p = 0.836<br>95% CI [-0.31, 0.38] |
| ODI and FWIT Interference (changes) | r(59) = -0.01<br>p = 0.934<br>95% CI [-0.27, 0.25] | r(28) = 0.09<br>p = 0.638<br>95% CI [-0.29, 0.45] | r(31) = -0.01<br>p = 0.940<br>95% CI [-0.37, 0.34] |
| ODI and Rey-Osterrieth memory (changes) | r(66) = -0.14<br>p = 0.251<br>95% CI [-0.37, 0.10] | r(34) = 0.04<br>p = 0.838<br>95% CI [-0.30, 0.37] | r(32) = -0.31<br>p = 0.081<br>95% CI [-0.60, 0.04] |

*Note.* CI: confidence interval; CON: control group; FWIT: Stroop interference test (German version); NDI: Neurite density index; ODI: orientation dispersion index; PAG: physical activity group.
